# Virtual rehabilitation for individuals with Long COVID: a randomized controlled trial

**DOI:** 10.1101/2024.11.24.24317856

**Authors:** Tania Janaudis-Ferreira, Marla K. Beauchamp, Amanda Rizk, Catherine M. Tansey, Maria Sedeno, Laura Barreto, Jean Bourbeau, Bryan A. Ross, Andrea Benedetti, Pei Zhi Li, Kriti Agarwal, Rebecca Zucco, Julie Lopez, Emily Crowley, Julie Cloutier

## Abstract

**Background:** Our primary objective was to investigate whether an 8-week virtual rehabilitation program for individuals with long COVID improves functional mobility compared to usual care.

**Methods:** Subjects were randomly assigned to receive either i) virtual rehabilitation plus usual outpatient care or ii) usual outpatient care. The intervention group underwent an 8-week virtual rehabilitation program which consisted of personalised and symptom-titrated functional aerobic and resistance exercises as well as long COVID educational sessions. The primary outcome was the Activity Measure for Post-Acute Care (AM-PAC) mobility score. Secondary outcomes included the Baseline and Transition Dyspnea Index (BDI/TDI), the Fatigue Visual Analog Scale, 12-item short-form, EuroQol 5 Dimension 5 Level (EQ-5D-5L), DePaul Symptom Questionnaire – PEM, physical function tests, questionnaires on mental health, acceptability and adverse events.

**Findings:** 132 individuals with long COVID (mean age 48 ± 11.8; 75% female) were enrolled. The adherence rate was 96%; however, 25 participants (39%) in the intervention group were unable to progress their exercises through the FITT (frequency, intensity, time, and type) principle due to symptoms. No between group differences were found for change in AM-PAC mobility (95% CI –0.91 to 2.13). The proportion of participants achieving the minimal detectable change in the AM-PAC mobility at the end of the intervention period was higher in the intervention group (35.8% (SE 6.0%) vs. 17.0% (SE 4.7%)) (95% CI 3.9 to 33.8). Compared with controls, scores on the EQ-5D-5L pain/discomfort (95% CI –0.70 to –0.03), EQ-5D-5L VAS (95% CI 1.05 to 14.43), VAS fatigue (95% CI –1.78 to –0.02), as well as for the TDI functional (95% CI 0.07 to 0.72), effort (95% CI 0.10 to 1.12) and total scores (95% CI 0.10 to 2.37) were greater in the intervention group. There were no between-group differences in other outcomes and no serious adverse events.

**Interpretation:** An 8-week virtual rehabilitation program did not improve self-reported mobility for most patients with long COVID, however we did find improvements in health status and symptom persistence. Progression of exercise training is challenging in this population.

## INTRODUCTION

Coronavirus disease (COVID-19), an infectious disease caused by SARS-CoV-2 virus, was responsible for the global pandemic declared in March 2020 by the World Health Organization (WHO).^1^ The COVID-19 infection can result in long-term health consequences, with symptoms persisting for months or even years following the initial diagnosis.^1^ Defined by the WHO as long COVID or Post COVID-19 condition, these symptoms emerge or persist three months after the initial SARS-CoV-2 infection, lasting for at least two months without an alternate explanation.

Globally, an estimated 148 million individuals are diagnosed with long COVID,^2^ with approximately 1.2 million affected in Canada^2^. Beyond individual health implications, the repercussions extend to societal and economic realms, with around 100,000 Canadian adults unable to resume work or schooling due to persisting symptoms.^3^

While fatigue, shortness of breath, memory and concentration problems are most commonly reported, long COVID encompasses a wide spectrum of over 200 possible manifestations,^4^ significantly impacting daily functioning.^4^ In a US nationwide survey study involving 7926 individuals with long COVID (median number of days between infection and survey completion was 325), 41% were classified as having mobility disability.^5^ In a Canadian prospective cohort study, 55.3% of 254 patients who had been hospitalized for COVID-19 infection demonstrated clinically important deficits in mobility and in performance-based tests of physical function at 12 months of follow-up.^6^ Data from the large population-based Canadian Longitudinal Study on Aging similarly demonstrated new onset mobility problems even in the absence of hospitalization.^7^ These data suggest that individuals with long COVID may benefit from a physical rehabilitation intervention to address their persisting symptoms and functional limitation. Rehabilitation is now recommended by the WHO^1^ and other international societies,^8,9^ albeit with relevant caveats.

Post-exertional malaise (PEM) is defined as “the worsening of symptoms following even minor physical or mental exertion, with symptoms typically worsening 12 to 48 hours after activity and lasting for days or even weeks”.^10^ Given the high proportion of PEM in individuals with long COVID,^11^ it is recommended that physical rehabilitation for individuals with long COVID offer a personalized program for those who feel prepared to increase their level of physical activity beyond their current daily activities and/or would like to incorporate exercise into managing their condition.^1,12^ Symptom-titrated exercise training is an appropriate strategy for individuals with long COVID as it keeps the exercise parameters within individualized symptom thresholds to avoid PEM while still promoting physical activity and functional improvement.^12^ Graded exercise training, commonly used in rehabilitation in different chronic diseases,^13^ involves making fixed incremental increases in exercise intensity over time, and is not recommended in long COVID as it may exacerbate symptoms or cause PEM.^1^

Although emerging evidence suggests that rehabilitation interventions for individuals with long COVID improve functional exercise capacity, dyspnea, and quality of life,^14^ several shortcomings exist in the few available trials.^14^ Most studies were limited to only previously hospitalized patients, and did not systematically monitor adverse events or addressed strategies to avoid PEM during exercise sessions.^14^ Furthermore, these trials often omitted critically important outcome measures such as mobility and fatigue, which are essential for evaluating the impact of COVID-19 on patient functions in daily life.^14^ Additionally, the majority of these trials provided rehabilitation programs exclusively in person.^14^ Virtual rehabilitation programs, previous to and after the pandemic, has been considered a promising avenue for delivering exercise-based therapies as they have the potential to enhance uptake and adherence.^15,16^

The primary objective of this study is to evaluate the effect of an 8-week virtual home-based rehabilitation program added to usual care on functional mobility in individuals with long COVID, compared to usual care. Secondary objectives are to evaluate the impact of the intervention on lower body strength; symptoms of fatigue and dyspnea; PEM, health-related-quality-of-life (HRQOL); anxiety, depression, distress, cognitive function; health service use and adverse events; and to assess participants’ satisfaction with the intervention.

## METHODS

This study is reported following the CONSORT guidelines for reporting randomized controlled trials.^17^

### Study Design

The present study used a prospective, multicenter, assessor-blind, randomized controlled trial (RCT) design. The study was conducted at the McGill University Health Centre in Montreal, Quebec and McMaster University in Hamilton, Ontario, Canada. The study was approved by the institutional ethics committees of the Research Institute of McGill University Health Centre (RI-MUHC) and McMaster University and was registered at ClinicalTrials.gov (identifier NCT05298878).

### Participants

Subjects were recruited from the MUHC COVID-19 clinic, MUHC COVID-19 Biobank, Quebec Action for Post-COVID Project, the McMaster Functional Recovery of Hospitalized Patients with COVID-19 Prospective Cohort Study^6^ and social media (posts targeting those living with long COVID in the provinces of Ontario and Quebec).

Inclusion criteria were as follows: (1) adult individuals (≥18 years old) with confirmed or probable COVID-19 infection with self-reported persisting symptoms of either reduced mobility, muscle weakness, dyspnea, or fatigue; (2) technologically capable of connecting (either independently or through household members) with an online video conferencing platform through an e-mail invitation; and (3) able to collaborate with the research assistant to complete the virtual assessment sessions or have a family member available to help. Exclusion criteria were: (1) pre-existing or newly identified severe cognitive impairment; (2) inability to speak or comprehend English or French; or (3) known or self-reported acute and/or uncontrolled cardiac, musculoskeletal, or neurological condition that might render rehabilitation participation unsafe. Participants provided written informed consent.

### Randomization and Masking

Eligible subjects who agreed to participate in the study were randomly allocated in a 1:1 ratio to receive either a virtual home-based rehabilitation program in addition to usual outpatient care (intervention group) or usual outpatient care alone (control group). The randomization sequence was created by the team biostatistician using computer-generated (Blockrand package in R), permuted, balanced blocks of varying sizes (2-8) and was stratified by recruitment location, either Quebec or Ontario. The randomization list was then exported to REDCap,^18^ an electronic data capture tool hosted at the MUHC. The principal investigator (TJF) randomized the participants after the baseline assessments and notified them of the group allocation. The principal investigator also notified the team of kinesiologists when a participant was randomized to the intervention group. The assessors involved in recruitment and data collection, as well as the data analyst, were blinded to the group allocation.

### Procedures

Participants underwent baseline evaluations, completed an 8-week rehabilitation protocol, underwent post-8-week evaluations, and provided self-reported healthcare service usage at 30 days after the evaluation. They also completed a questionnaire about demographic information (age, sex, gender, height, weight, education level, household income, work status, smoking history, comorbidities, hospital and intensive care unit length of stay, number of COVID-19 infections, medication, supplemental oxygen use and level of physical activity). The protocol (exercise training and assessments) was 100% virtual using the Zoom for Healthcare Platform. The performance-based tests and the primary outcome (The Activity Measure for Post-Acute Care (AM-PAC))^19^ were conducted at baseline and at post-8-week evaluations. Questionnaires were collected using a REDCap link^18^ that was sent to participants.

### Usual Care (Control group)

Subjects in the usual care group received usual outpatient care which consisted of any outpatient visits with a physician or other healthcare professional. At the study entry, they received a PDF document prepared by the research team containing generic written instructions on how to manage symptoms and safely engage in physical activity while avoiding the “push and crash” cycle that may occur with individuals with PEM.^12^

### Virtual Rehabilitation Program (Intervention Group)

Participants in the intervention group underwent an 8-week virtual home-based rehabilitation program. The program delivery was executed by bilingual, trained, and certified kinesiologists hired in partnership with the Willkin Health organization (https://willkin.ca/en/about-us/).

During the first supervised session, the kinesiologist discussed safety of remote sessions and goal setting, chose individualized exercises, and established rapport with the subject. The intervention included one-on-one sessions with kinesiologists including a phase-out design to encourage autonomy in the long-term. During Weeks 1-2, subjects were asked to engage in three supervised Zoom sessions/week. In Weeks 3-4, they participated in two supervised Zoom sessions/week and 1 independent session/week. Weeks 5-8 involved one supervised Zoom session/week and 2 independent sessions/week. The goal of each virtual supervised session with the kinesiologist was to last 40 minutes and include a combination of education, breathing exercises, aerobic, functional lower and upper body strengthening and flexibility exercises (Table 1 – supplemental). Heart rate and oxygen saturation were monitored during each session using an oximeter provided to participants before baseline assessments. The kinesiologist asked questions regarding dyspnea level using the Borg 0-10 scale, and rate of perceived exertion (RPE) using the Borg 6-20 scale at the beginning and end of each supervised session. At the beginning of each session, the kinesiologist asked two questions to screen for worsening in symptoms: “did you experience a worsening of your fatigue/energy levels after engaging in the last exercise session” and “did you experience any change in symptoms”.

**Table 1.** Baseline Characteristics.

|  | **Total**<br>**(n = 132)** | **Control**<br>**Group**<br>**(n = 67)** | **Intervention**<br>**Group**<br>**(n = 65)** | **Drop-outs**<br>**(n = 5)** |
| --- | --- | --- | --- | --- |
| **Site, n (%)** |  |  |  |  |
| MUHC | 114 (86.4) | 57 (85.1) | 57 (87.7) | 4 (80.0) |
| McMaster | 18 (13.6) | 10 (14.9) | 8 (12.3) | 1 (20.0) |
| **Age (years)** | 48.0 ± 11.8 | 48.0 ± 11.7 | 48.0 ± 12.0 | 48.6 ± 14.1 |
| **Sex, n (%)** |  |  |  |  |
| Female | 99 (75.0) | 53 (79.1) | 46 (70.8) | 4 (80.0) |
| Male | 33 (25.0) | 14 (20.9) | 19 (29.2) | 1 (20.0) |
| **Gender, n (%)** |  |  |  |  |
| Woman | 97 (73.5) | 52 (77.6) | 45 (69.2) | 4 (80.0) |
| Man | 33 (25.0) | 14 (20.9) | 19 (29.2) | 1 (20.0) |
| Gender queer/Gender non-conforming | 2 (1.5) | 1 (1.5) | 1 (1.5) | 0 (0.0) |
| **Ethnicity, n (%)** |  |  |  |  |
| Asian | 4 (3.1) | 1 (1.5) | 3 (4.8) | 0 (0.0) |
| Black | 1 (0.8) | 1 (1.5) | 0 (0.0) | 0 (0.0) |
| Caucasian | 111 (86.7) | 56 (84.8) | 55 (88.7) | 4 (80.0) |
| Latino American | 1 (0.8) | 1 (1.5) | 0 (0.0) | 0 (0.0) |
| Middle East/North African | 3 (2.3) | 0 (0.0) | 3 (4.8) | 1 (20.0) |
| Other | 8 (6.3) | 7 (10.6) | 1 (1.6) | 0 (0.0) |
| **BMI, kg/m<sup>2</sup> (self-reported)** | 31.3 ± 17.0 | 33.4 ± 22.2 | 29.2 ± 9.2 | 29.9 ± 17.7 |
| **Education Highest degree attained, n (%)** |  |  |  |  |
| High school | 16 (12.1) | 7 (10.4) | 9 (13.8) | 0 (0.0) |
| Undergraduate | 42 (31.8) | 20 (29.9) | 22 (33.8) | 0 (0.0) |
| College | 31 (23.5) | 16 (23.9) | 15 (23.1) | 2 (40.0) |
| Graduate | 36 (27.3) | 19 (28.4) | 17 (26.2) | 3 (60.0) |
| Other | 7 (5.3) | 5 (7.5) | 2 (3.1) | 0 (0.0) |
| **Household income, n (%)** |  |  |  |  |
| Less than \$25,000 | 6 (5.4) | 4 (6.9) | 2 (3.8) | 0 (0.0) |
| \$25,000 to \$50,000 | 11 (9.9) | 4 (6.9) | 7 (13.2) | 1 (20.0) |
| \$50,001 to \$75,000 | 23 (20.7) | 9 (15.5) | 14 (26.4) | 1 (20.0) |
| \$75,001 to \$100,000 | 24 (21.6) | 13 (22.4) | 11 (20.8) | 0 (0.0) |
| \$100,001 or more | 47 (42.3) | 28 (48.3) | 19 (35.8) | 3 (60.0) |
| **Work status, n (%)** |  |  |  |  |
| Employed/self-employed | 95 (72.0) | 51 (76.1) | 44 (67.7) | 2 (40.0) |
| Employed/self-employed yet on leave | 77 (81.1) | 40 (78.4) | 37 (84.1) | 1 (50.0) |
| Unemployed | 13 (9.8) | 7 (10.4) | 6 (9.2) | 1 (20.0) |
| Studying | 2 (1.5) | 1 (1.5) | 1 (1.5) | 0 (0.0) |
| Retired | 10 (7.6) | 2 (3.0) | 8 (12.3) | 1 (20.0) |
| Other | 12 (9·1) | 6 (9·0) | 6 (9·2) | 1 (20·0) |
| **Cigarette smoking status, n (%)** |  |  |  |  |
| Current smoker | 0 (0·0) | 0 (0·0) | 0 (0·0) | 0 (0·0) |
| Previous smoker | 51 (38·6) | 26 (38·8) | 25 (38·5) | 2 (40·0) |
| Never smoked | 81 (61·4) | 41 (61·2) | 40 (61·5) | 3 (60·0) |
| **Comorbidities, n (%)** |  |  |  |  |
| Hypertension | 24 (18·2) | 12 (17·9) | 12 (18·5) | 1 (20·0) |
| Diabetes | 10 (7·6) | 6 (9·0) | 4 (6·2) | 1 (20·0) |
| Musculoskeletal disease | 30 (22·7) | 17 (25·4) | 13 (20·0) | 1 (20·0) |
| Cardiac disease | 8 (6·1) | 4 (6·0) | 4 (6·2) | 0 (0·0) |
| Cancer | 0 (0·0) | 0 (0·0) | 0 (0·0) | 0 (0·0) |
| Respiratory disease (other than COVID) | 19 (14·4) | 8 (11·9) | 11 (16·9) | 2 (40·0) |
| Number of comorbidities, median (Q1, Q3) | 1·0 (0·0, 2·0) | 1·0 (0·0, 2·0) | 0·0 (0·0, 1·0) | 1·0 (0·0, 3·0) |
| Number of comorbidities as category |  |  |  |  |
| 0 | 63 (47·7) | 30 (44·8) | 33 (50·8) | 2 (40·0) |
| 1 | 34 (25·8) | 18 (26·9) | 16 (24·6) | 1 (20·0) |
| 2+ | 35 (26·5) | 19 (28·4) | 16 (24·6) | 2 (40·0) |
| **Taking prescribed medication, n (%)** | 113 (85·6) | 58 (86·6) | 55 (84·6) | 4 (80·0) |
| **COVID Frequency, median (Q1, Q3)** | 1·0 (1·0, 2·0) | 1·0 (1·0, 2·0) | 1·0 (1·0, 2·0) | 1·0 (1·0, 1·0) |
| **Long covid duration, in months, median (Q1, Q3)** | 13·7 (8·4, 21·3) | 13·7 (9·2, 21·3) | 13·6 (7·8, 21·1) | 14·6 (9·5, 18·6) |
| **Number of symptoms, median (Q1, Q3)** | 4·0 (3·0, 6·0) | 4·0 (3·0, 6·0) | 5·0 (3·0, 7·0) | 5·0 (4·0, 6·0) |
| **Hospitalization** |  |  |  |  |
| Hospitalized due to COVID, n (%) |  |  |  |  |
| 0 | 103 (78·0) | 52 (77·6) | 51 (78·5) | 4 (80·0) |
| 1 | 25 (18·9) | 14 (20·9) | 11 (16·9) | 1 (20·0) |
| 2+ | 4 (3·0) | 1 (1·5) | 3 (4·6) | 0 (0·0) |
| 1st Hospitalization LOS | 7·2 ± 32·5 | 10·3 ± 42·6 | 4·0 ± 16·2 | 2·8 ± 6·3 |
| ≥1 ICU admission for COVID, n (%) | 15 (11·4) | 9 (13·4) | 6 (9·2) | 0 (0·0) |
| 1st ICU LOS | 4·8 ± 26·7 | 7·7 ± 36·7 | 1·9 ± 7·4 | 0·0 ± 0·0 |
| **Number of vaccinations, median (Q1, Q3)** | 4·0 (3·0, 4·0) | 4·0 (3·0, 4·0) | 3·0 (2·0, 4·0) | - |
| **Supplemental oxygen (O<sub>2</sub>), n (%)** | 2 (1·5) | 2 (3·0) | 0 (0·0) | 0 (0·0) |
| **Current level of activity, n (%)** |  |  |  |  |
| Vigorously active | 0 (0·0) | 0 (0·0) | 0 (0·0) | 0 (0·0) |
| Moderately active | 29 (22·0) | 14 (20·9) | 15 (23·1) | 1 (20·0) |
| Seldom active | 103 (78·0) | 53 (79·1) | 50 (76·9) | 4 (80·0) |
| **BDI Total score (0 - 12), median (Q1, Q3)** | 3·5 (2·0, 5·0) | 3·0 (2·0, 5·0) | 4·0 (2·0, 5·0) | 3·0 (3·0, 4·0) |
| **DSQ-PEM** |  |  |  |  |
| PEM (sum of all scores) (0 - 5), median (Q1, Q3) | 4·0 (3·0, 5·0) | 4·0 (3·0, 5·0) | 4·0 (3·0, 5·0) | 4·0 (3·0, 5·0) |
| PEM score, n (%) |  |  |  |  |
| 0 | 9 (6·8) | 7 (10·4) | 2 (3·1) | 0 (0·0) |
| 1-2 | 17 (12·9) | 4 (6·0) | 13 (20·0) | 1 (20·0) |
| 3-5 | 106 (80·3) | 56 (83·6) | 50 (76·9) | 4 (80·0) |
| PEM present with minimal physical efforts, n (%) | 119 (90·2) | 60 (89·6) | 59 (90·8) | 5 (100·0) |
| PEM present with mental effort, n (%) | 106 (80·3) | 51 (76·1) | 55 (84·6) | 4 (80·0) |

Data are presented as mean ± SD unless otherwise specified
MUHC: McGill University Health Centre; BMI: body mass index; LOS: length of stay; ICU: intensive care unit; BDI: Beck Depression Inventory; DSQ-PEM: DePaul Symptom Questionnaire - post-exertional malaise.

The target intensity for aerobic and strengthening exercises was moderate (12-14 points on Borg 6-20 scale). However, initial intensities varied by participant; for example, we targeted Borg 9-11/20 (light intensity) for severely deconditioned participants or those who reported PEM at study entry. Training progression was tailored to each subject and, when deemed safe by the kinesiologists, included modifications to frequency, intensity, type of exercise, or a combination of these factors. These progressive modifications were guided by participant feedback (e.g., improvements on the Borg 6-20 with no symptoms of PEM) at the beginning and end of each session (Table 2 – supplemental). Exercise regression was also considered in case of considerable worsening in Borg RPE scale after the session, or if participants reported PEM as an adverse event related to the intervention. During the independent sessions, participants performed the same exercises they were performing during the virtual sessions on the given week.

**Table 2.**
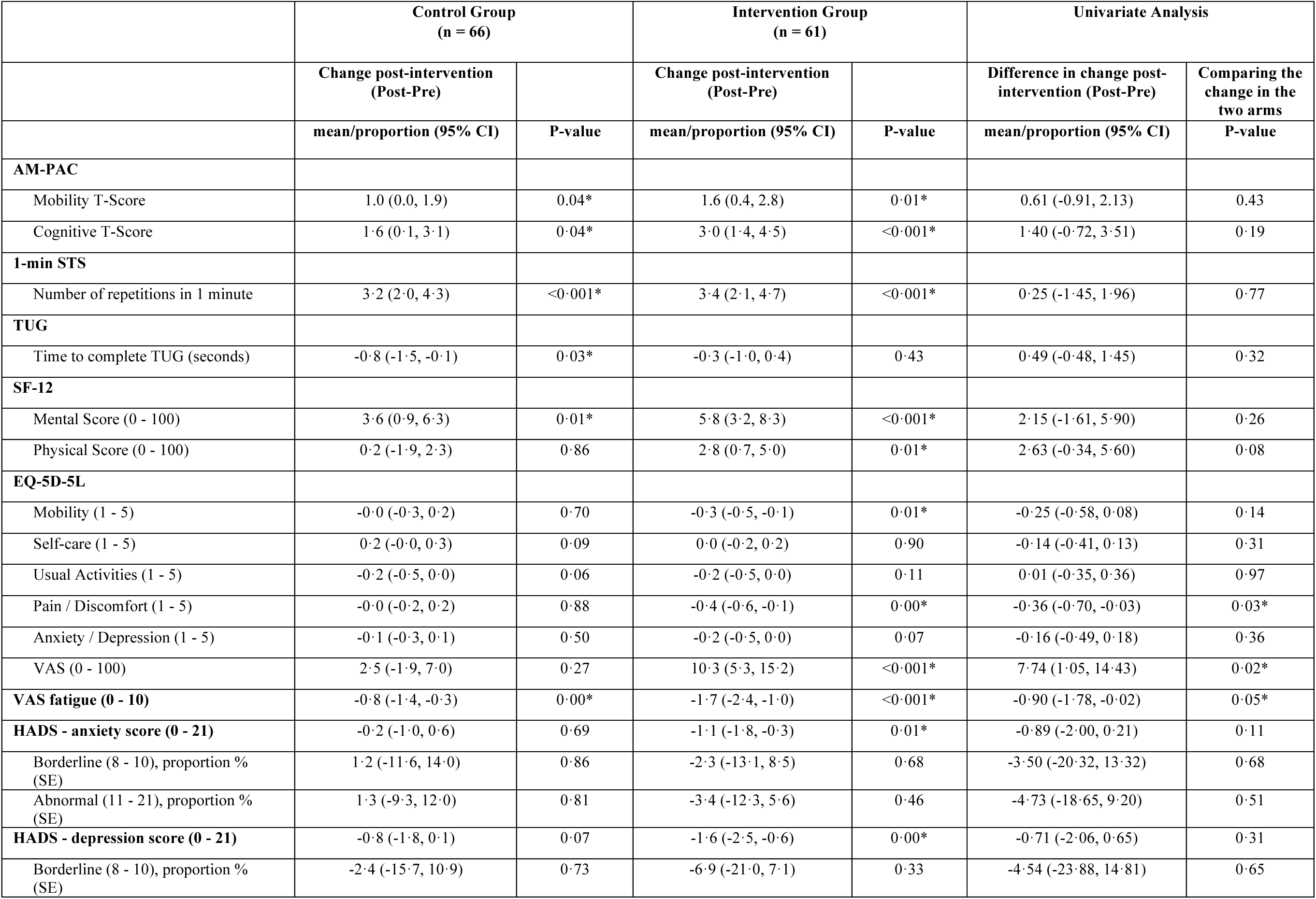
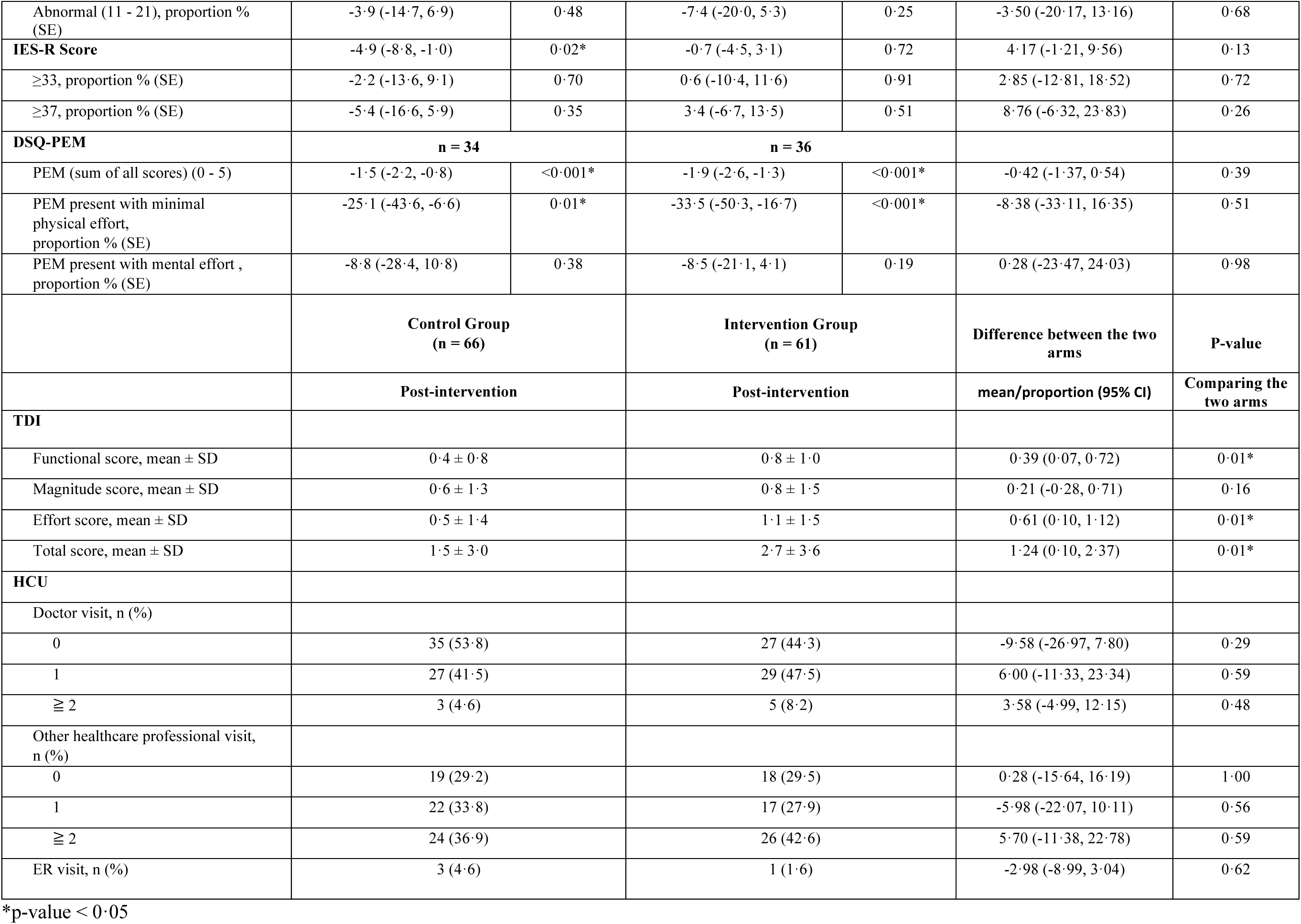
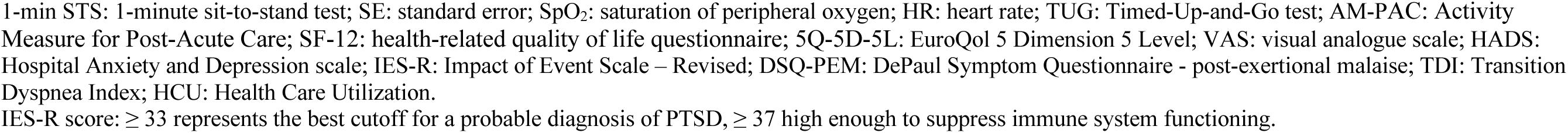
Between-Group Analyses.

The educational sessions were delivered once a week at the beginning of the first supervised Zoom session of the week. The educational materials were based on the “Living Well Beyond COVID-19” module (available free of charge at https://chroniclungdiseases.com/en/resources/covid-19/) developed in collaboration with RESPIPLUS (https://respiplus.com), an established non-profit organization and leader in developing self-management educational materials for chronic lung disease in Canada and internationally.^20^ These sessions covered various topics, including respiratory care (e.g., coping with dyspnea during activities of daily living and cough techniques), fatigue management (e.g., PEM, pacing and energy conservation), posture and injury prevention, nutritional management, psychological care (stress and anxiety), and social issues (e.g., return to work, social isolation). Participants in the intervention group received the same PDF document that was sent to the control group which included generic instructions on how to manage symptoms and safely engage in physical activity.

### Fidelity of the Intervention

Kinesiologists involved in the study took an online training program “Implementing a virtual home-based exercise program for individuals with long COVID” prepared by the research team. The course included educational information on physical rehabilitation in long COVID, quizzes to check participant understanding, as well as details on the study, reporting, and exercise training protocols. The course was offered via www.expandcourses.com before the study started to ensure that the intervention would be delivered as intended. Additionally, Willkin Health run bi-weekly meetings with the kinesiologists to ensure quality control.

### Outcome Measures

#### Primary Outcome Measure

The primary outcome was a change in the basic mobility domain of the Activity Measure for Post-Acute Care (AM-PAC).^19^ The AM-PAC is a patient-reported activity limitation instrument based on the International Classification of Functioning, Disability, and Health (ICF)^21^ that assesses 3 domains: basic mobility, daily activities, and applied cognition. Each item is scored from 1 (unable to perform) to 4 (none or no difficulty) with lower scores indicating lower levels of function. The AM-PAC has been validated for patients receiving post-acute care services and shown to be more responsive to change than the often-used Functional Independence Measure (FIM).^22^ The minimal detectable change (MDC) for the AM-PAC basic mobility is 3.3 and the minimal clinically important difference (MCID) based on distribution-based methods ranges between 1.7 and 4.2.^23,24^

#### Secondary Outcome Measures

##### Clinical Outcomes

The following outcome measures were assessed at baseline and after the intervention period (at 8-weeks post-baseline): 1-minute sit-to-stand test (1-min STS),^25^ the Fast Timed-Up-and-Go test (TUG),^26^ the Baseline and Transition Dyspnea Index (BDI/TDI),^27^ the Fatigue Visual Analog Scale,^28^ 12-item short-form (SF-12), EuroQol 5 Dimension 5 Level (EQ-5D-5L),^29^ DePaul Symptom Questionnaire – PEM (DSQ-PEM),^30^ AM-PAC cognition subscale,^19^ Hospital Anxiety and Depression scale (HADS),^31^ Impact of Event Scale – Revised (IES-R).^32^

At thirty days after the 8-week period, subjects provided information on self-reported healthcare service use in the past 30 days, including doctor visits, visits to other healthcare professionals, emergency department visits, and hospital admissions.

##### Adverse Events

Adverse events were monitored and categorized as “definitely related”, “possibly related”, or “unrelated” to the intervention.^33^ Furthermore, these events were categorized by category (musculoskeletal, cardiovascular, respiratory, multisystem.^33^ The severity of an adverse event was graded on a scale, with classifications ranging from Grade 1 for mild events (mild symptoms, medication intervention not indicated), Grade 2 for moderate (minimal local or non-invasive intervention indicated; limiting age-appropriate instrumental activities of daily living), Grade 3 for severe (severe or medically significant but not immediately life-threatening; hospitalization indicated), Grade 4 for life-threatening, and Grade 5 for death.^33^

##### Adherence rates

Adherence rates were calculated for each participant in the intervention group. The kinesiologist noted the attendance in the supervised sessions and participants were asked if they completed their independent session(s) at the beginning of each supervised session. Intervention adherence was defined as completion of ≥80% of all scheduled virtual and independent sessions.

##### Satisfaction with the Intervention

A short 17-question survey was sent to participants who were randomized to the intervention group. Questions 1 to 5 were on a semantic differential scale ranging from 1 to 5 and questions 6-17 were on a 1 to 7-point Likert scale.^34^ The questionnaire collected information on participants satisfaction with the structure of the program (number of sessions, length of session etc.), ease of use of the virtual platform, confidence with the exercises and utility for other patients with long COVID. The final question was open-ended, inviting participants to suggest improvements to the program.

### Statistical Analyses

Analyses were performed according to the intention-to-treat principle at the end of the study (8-week period + 30 days to collect self-reported healthcare utilization). Baseline data are presented as means ± standard deviation unless otherwise specified. Multiple imputation via chained equations was employed, with baseline age, sex, BMI, and pre-test values were included in the model to do the imputation and 10 imputed data sets. Generalized Estimating Equation (GEE) models were used to estimate within-group changes, with normal distribution via an identity link for continuous variables, and binary distribution with identity link function for categorical variables to estimate proportion difference. Between-group differences in the changes were tested for the pre-post-by-arms interactions to estimate whether the average change in the measures from pre to post differed in the two arms. A repeated statement was included in all models to account for possible correlation within subjects. In adjusted analyses, we fitted GEE models and adjusted for age and number of comorbidities which are known to be strongly correlated with mobility. If the GEE models did not converge for the correlated binary outcomes, a logistic regression model were used to estimate the proportion difference. We also conducted a per-protocol analysis that only included subjects in the intervention group who were able to progress the exercise training according to at least one of the following criteria: intensity, duration, time or type of exercise, and adjusted for important covariates. Sex-stratified analyses (separate models for males and females) were performed, and the effect modification of sex was tested using the interaction term pre-post-by-sex. Statistical significance was defined as two-sided p-value < 0.05. Statistical analyses were conducted using the SAS version 9.4 software.

The sample size calculation was performed using Proc power in SAS for a two groups simple T-test with a randomization 1:1. We assumed that the average difference in the change of AM-PAC from baseline to 8-weeks would be 3.3 points (the MDC), and the SD for each group to be equal to 5.0.^23,24^ When adjustments for covariates are needed, the estimated sample size is adjusted by a multiplier based on the strength of potential confounding of the covariates. Therefore, we will need a total of 132 subject (66 in the intervention group and 66 in the control group) to enable adjustment for four potential covariates for a 2-tailed test with a power of 90%, an alpha error of 0.05, and an estimated attrition rate of 10%.^35^

## RESULTS

Recruitment occurred between August 1^st^ 2022 and July 7^th^ 2023. Of the 301 individuals considered for screening, 132 individuals with long COVID were randomized to either the intervention group (n=65) or control group (n=67) (Figure 1). One hundred twenty-seven completed the post-assessment and were included in the analysis, leading to a retention rate of 96% (5 dropouts) (Figure 1).

**Figure 1.**
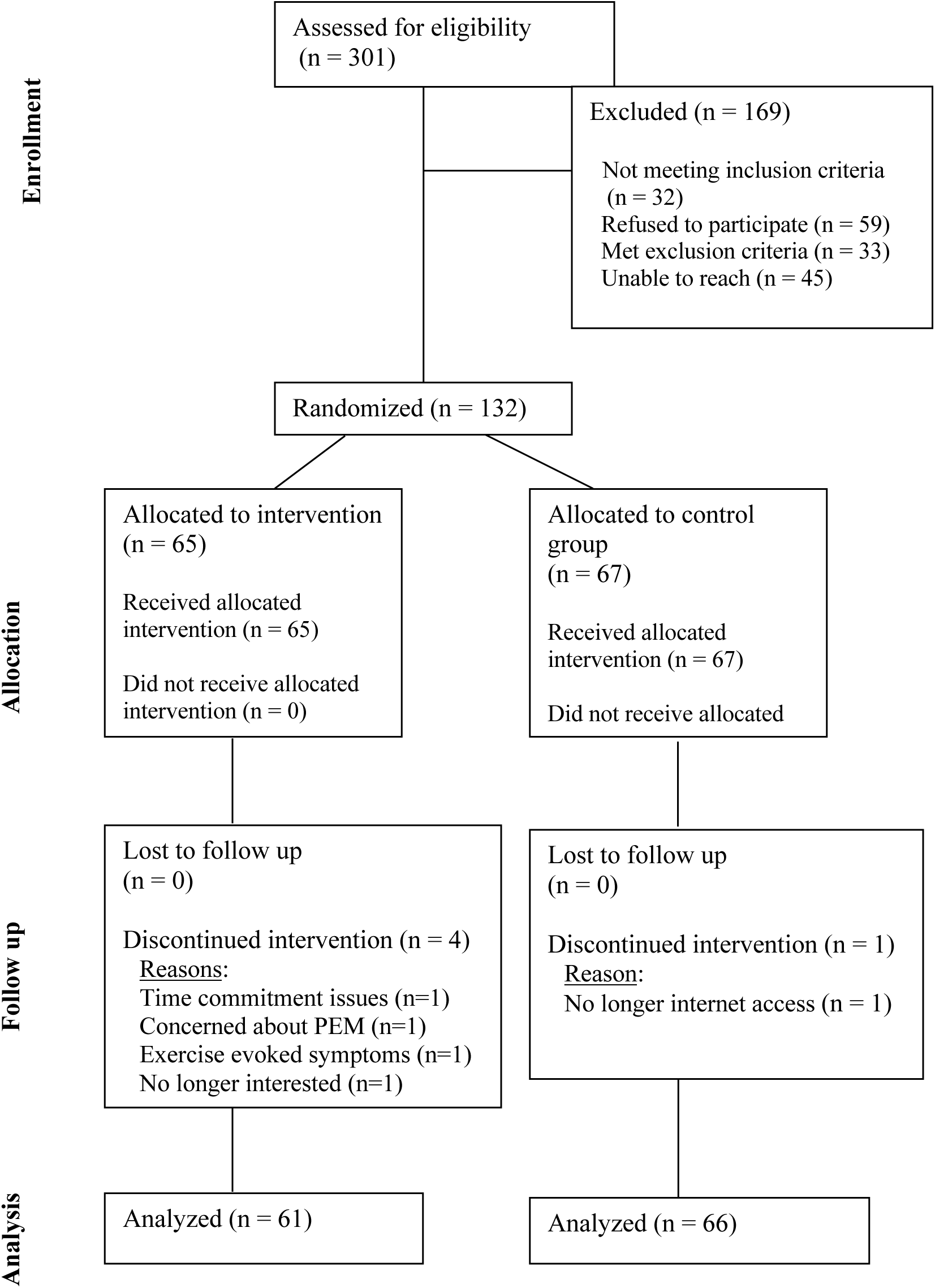
Study Flowchart

Baseline characteristics are presented in Table 1. Our sample was predominantly female (75%) with a mean age of 48 years (SD 11.8) and long COVID symptom duration of 13.7 months (IQR 8.4 – 21.3). The majority of the sample had a post-secondary degree and were employed or self-employed (72%), 81.1 % of which were on leave. Over 42.3% had a household income higher than $100,000 $CAN. The median number of comorbidities was 1(IQR 0 – 2.0). The majority of the sample had not been hospitalized due to COVID-19 infection (78%), had a median number of symptoms of 4, were vaccinated (median of 4 times), were seldom active at the time of completing the questionnaire (78%) and had experienced symptoms of PEM at the study entry (93.2%) (Table 1).

Participants in the intervention group attended a mean of 13.4 supervised exercise sessions out of a total possible of 14 sessions (adherence rate of supervised sessions of 96%) and a mean of 7.4 independent sessions out of a total possible of 10 sessions (adherence rate of independent sessions of 74%). Twenty-five participants (39%) in the intervention group could only perform simple range of motion and flexibility exercises or were unable to progress their exercises through the FITT (frequency, intensity, time and type) principle due to symptoms (e.g., dyspnea, fatigue, pain or headache).

No between group differences were found for change in AM-PAC mobility, the primary outcome (95% CI –0.91 to 2.13) (Table 2). Results from analyses that adjusted for age, sex, BMI, number of comorbidities and smoking status were similar to the results in the unadjusted analyses for AM-PAC mobility. The proportion of participants achieving the MDC of 3.3 in the AM-PAC mobility total score at the end of the intervention period, however, was higher in the intervention group (35.8% (standard error (SE) 6.0%) vs. 17.0% (SE 4.7%)) (95% CI 3.9 to 33.8) (P= .013)

Compared with the changes observed in the control group, in the unadjusted analyses, the magnitude of improvement in the intervention group was greater for EQ-5D-5L pain/discomfort (95% CI –0.70 to –0.03), EQ-5D-5L VAS (95% CI 1.05 to 14.43), VAS fatigue (95% CI –1.78 to – 0.02), as well as for the TDI functional (95% CI 0.07 to 0.72), effort (95% CI 0.10 to 1.12) and total scores (95% CI 0.10 to 2.37) (Table 2). There were no between-group differences in healthcare utilization at 30 days after the 8-week intervention period (Table 2). Results from analyses that adjusted for age, sex, BMI, number of comorbidities and smoking status were similar to the results in the unadjusted analyses for secondary outcomes.

No adverse events related to the intervention were classified as Grade 3 (severe), 4 (life-threatening), or 5 (death) (Table 3). The adverse events classified as “definitely” or “possibly” related to the intervention (45 adverse events reported by 30 participants)) were either Grade 1 (mild symptoms, medication intervention not indicated) (13 adverse events)) or Grade 2 (moderate symptoms, minimal local intervention required; limiting age-appropriate instrumental activities of daily living) (32 adverse events)) (Table 3). The most common types of adverse events were musculoskeletal and multisystem (Table 3). Twenty-eight adverse events were classified as PEM-related, with 11 of these “definitely related” and 17 “possibly related” to the intervention. All PEM-related adverse events were classified as either mild or moderate. Detailed data on the 36 adverse events categorized as “unrelated” to the intervention are available in Table 3 – supplement.

**Table 3.**
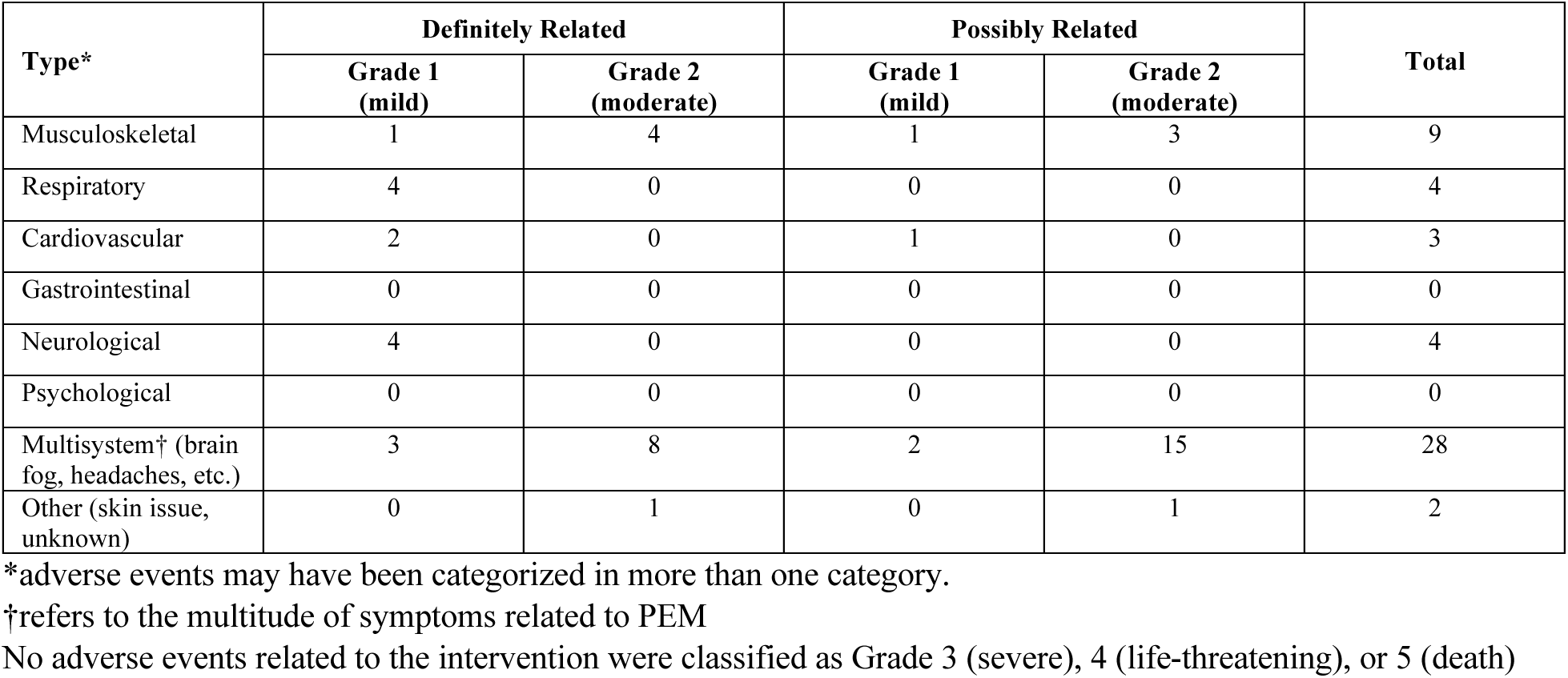
Types of Adverse Events Related to the Intervention.

| Type* | Definitely Related |  | Possibly Related |  | Total |
| --- | --- | --- | --- | --- | --- |
|  | Grade 1<br>(mild) | Grade 2<br>(moderate) | Grade 1<br>(mild) | Grade 2<br>(moderate) |  |
| Musculoskeletal | 1 | 4 | 1 | 3 | 9 |
| Respiratory | 4 | 0 | 0 | 0 | 4 |
| Cardiovascular | 2 | 0 | 1 | 0 | 3 |
| Gastrointestinal | 0 | 0 | 0 | 0 | 0 |
| Neurological | 4 | 0 | 0 | 0 | 4 |
| Psychological | 0 | 0 | 0 | 0 | 0 |
| Multisystem† (brain fog, headaches, etc.) | 3 | 8 | 2 | 15 | 28 |
| Other (skin issue, unknown) | 0 | 1 | 0 | 1 | 2 |

\*adverse events may have been categorized in more than one category.
†refers to the multitude of symptoms related to PEM
No adverse events related to the intervention were classified as Grade 3 (severe), 4 (life-threatening), or 5 (death)

In ad hoc analyses, when comparing the demographics of participants in the intervention group who did not experience adverse events (n = 35) with those who did (n = 30), those who experienced adverse events had a significantly higher proportion of comorbid cardiac disease (95% CI 11.70 to 25.50) and higher PEM scores (95% CI 0.26 to 1.76) (Table 4 – supplement). Furthermore, when comparing participants who did not experience adverse events (n = 35) with those who had PEM-related adverse events classified as “definitely” or “possibly” related (n = 22), the latter group had a significantly higher proportion of comorbid cardiac disease (95% CI 2.06 to 34.30), a shorter length of stay during their first hospitalization (95% CI –11.23 to –0.23), higher PEM scores at baseline (95% CI 0.11 to 1.55), and a greater proportion of participants with PEM triggered by mental effort (95% CI 8.96 to 36.77) (Table 5 – supplement).

**Table 4.**
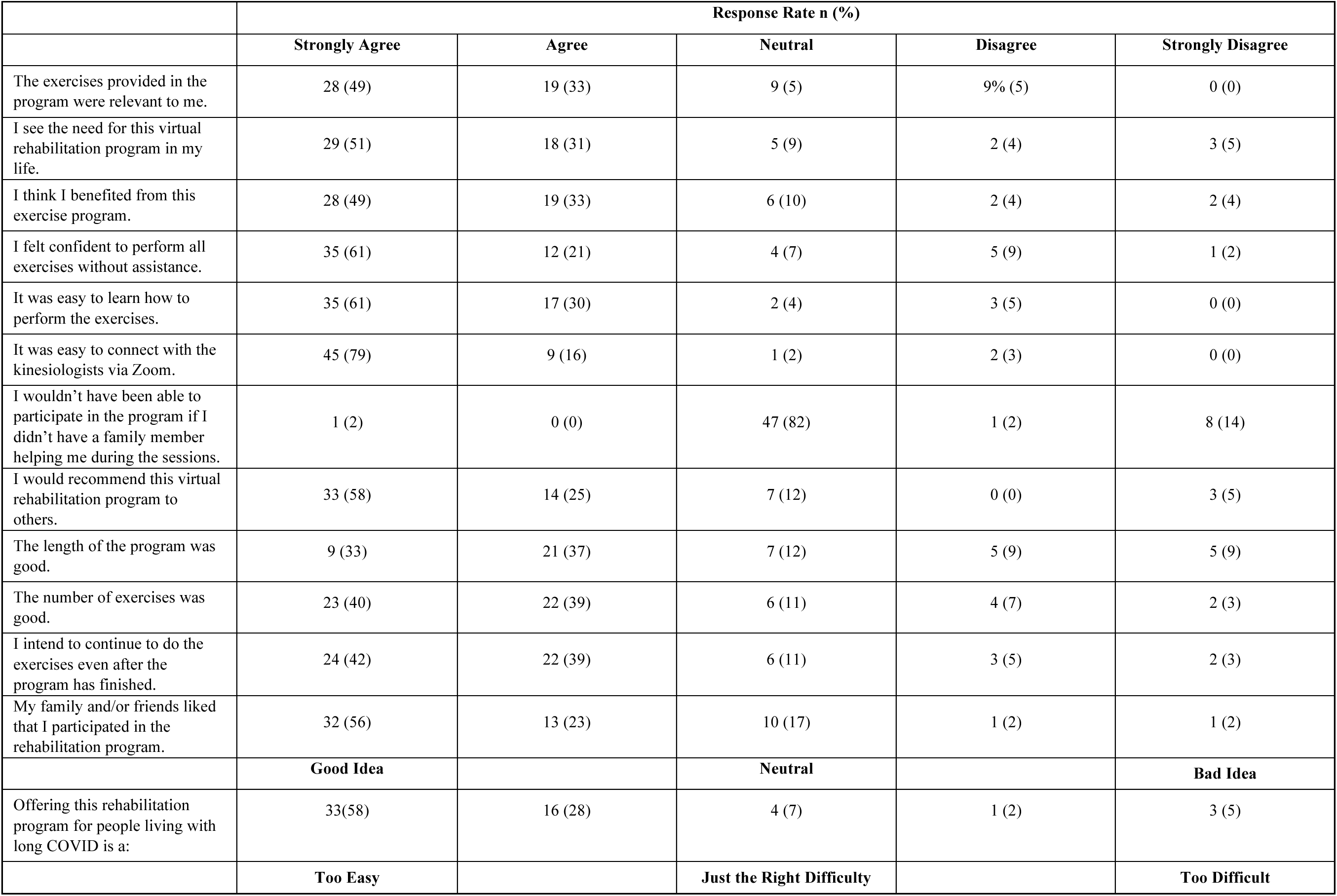
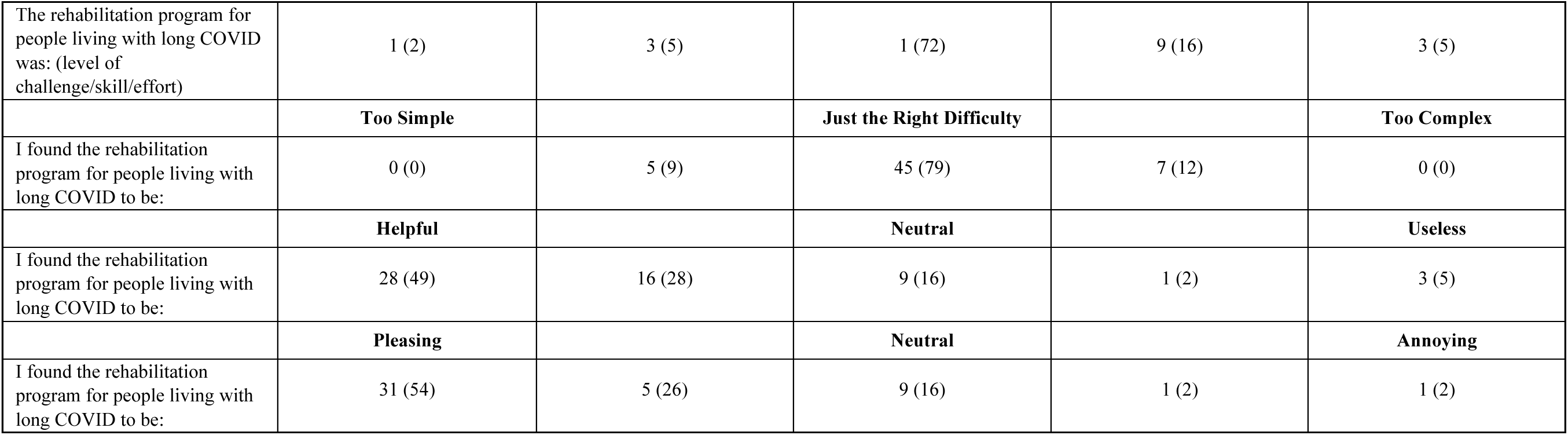
Participant Satisfaction with Intervention Program (57 respondents)

There were no statistically significant differences in baseline characteristics between the control group and the group of participants who were able to progress through the exercise program (n = 40) (Table 6 –supplement). Per-protocol analysis comparing the control group (n = 66) to participants in the intervention group who were able to progress through the exercise program (n = 36) showed improvements in SF-12 physical score (95% CI 0.61 to 7.46), EQ-5D-5L pain/discomfort (95% CI –0.84 to –0.06), EQ-5D-5L VAS (95% CI 3.24 to 19.73), VAS fatigue (95% CI –2.42 to –0.39), HADS anxiety score (95% CI –2.68 to –0.08) as well as in the TDI functional (95% CI 0.32 to 1.02), effort (95% CI 0.51 to 1.59), and total scores (95% CI 0.92 to 3.31) in favor of the intervention (Table 7 – supplement).

The changes in outcomes within the intervention and control groups were similar between male and female participants, except for the EQ-5D-5L VAS (greater improvement in females). (Table 8 – supplement).

Fifty-seven of 61 participants who completed the rehabilitation program filled out the satisfaction survey (response rate 93%; 70% female). Most respondents (83%) would recommend the program. The majority of the respondents reported that they either agreed or strongly agreed that the program was easy to learn (91%) and relevant to their needs (82%). Seventy-two percent of the respondents thought the program had just the right difficulty and 80% reported that the program was pleasant (Table 4). Positive free-text comments emphasized life-changing experiences such as “this program has changed my life” and “the program made me alive again”. Eight individuals (14%) commented negatively on how the program affected their symptoms, for example, “after most sessions, I was unable to be productive later in the day”.

Four respondents (7%) found the program not suitable. There were suggestions for a longer program with more time in between sessions to allow for extended recovery periods and a multidisciplinary approach.

## DISCUSSION

The primary objective of this study was to evaluate the effect of an 8-week virtual home-based rehabilitation program in addition to usual care on functional mobility in individuals with long COVID, compared to usual care alone. While there was no statistically significant between-group difference in our primary outcome, AM-PAC mobility, the proportion of participants achieving the MDC of 3.3 was twice as high in the intervention group compared to the control group. The intervention group also showed significant improvements in key secondary outcomes including fatigue, dyspnea, pain and discomfort, and perceived health with the virtual rehabilitation program. Our findings revealed high adherence rates, no severe adverse events, and the majority of participants expressed high satisfaction with the virtual rehabilitation program.

While the published MDC for AM-PAC mobility is not specific to the long COVID population, data from a long COVID cohort study suggest a MDC of 4.2.^6^ When we conducted the same analysis using the suggested MDC for long COVID, the findings remained consistent: the proportion of participants achieving the MDC of 4.2 was higher in the intervention group (23.4%) compared to the control group (14%) although not statistically significant different. It is possible that a disease-specific functional scale for COVID-19 would have been more sensitive to assess change following the intervention for this population or that another domain of functioning, besides mobility, would have shown greater benefit given the limitations in exercise progression observed in this trial. There were no significant between-group differences in the TUG and 1-min STS tests. This outcome may be attributed to the intervention group participants’ ability to pace themselves during physical activity, a strategy taught by the kinesiologists during the rehabilitation program. This enhanced pacing strategy might have obscured performance improvements in the intervention group.

An interesting and unexpected finding was that 39% of the participants in the intervention group could only perform simple range of motion and flexibility exercises or were unable to progress their exercises through the FITT principle due to symptoms such as fatigue, dyspnea, pain, and headache. This reflects not only the level of disability of the participants but also their ability to communicate their symptoms to the kinesiologists and, importantly, the kinesiologists’ careful consideration in recognizing when a regression of the exercise training or no exercise was warranted. It is worth noting that participants in the intervention group received education about PEM, which included information on how to recognize and mitigate it. The education component was essential in helping participants avoid exerting themselves beyond their limits.

While 52% of participants in the intervention group reported at least one adverse event that was definitely or possibly related to the intervention, these events were classified as mild or moderate, involving either minor symptoms that did not require medication, or limited disruptions to age-appropriate instrumental activities of daily living, which required only minimal local intervention (rest or medication as-needed). It is important to acknowledge that there was no monitoring of adverse events in the control group, which may have led to overreporting in the intervention group due to the careful monitoring process. Moreover, the fluctuating nature of long COVID makes classifying adverse events challenging, as recurrent symptoms may have been classified as adverse events. Notably, many adverse events were related to other activities participants engaged in during the intervention period such as hosting family/social events, taking care of pets, work with the computer and doing outdoor physical activities. A review and meta-analysis by Pouliopoulou et al. on rehabilitation interventions in long COVID, which included 14 trials involving 1244 patients with long COVID (8 of which included an exercise component with 429 patients) found that only 2 out of 8 trials reported adverse events, concluding that there is a high level of uncertainty and imprecision regarding adverse events in this population.^14^ An RCT by Espinoza-Bravo et al., which was not included in this meta-analysis, reported no adverse events in 43 participants randomized to either resistance training or aerobic exercise.^36^ Conversely, the RCT by McGregor et al.,^37^ that included 585 individuals with long COVID who had been hospitalized to participate in virtual exercises sessions, reported a total of 44 adverse events (16 in the usual care group and 28 in the intervention group), with 2 adverse events considered “definitely related” and 2 “probably related” to the intervention. They also reported 21 serious adverse events (7 in the usual care group and 14 in the intervention group), with 1 event considered “possibly related” to the intervention.^37^ The discrepancy between study findings may be attributed to several factors, including differences in exercise protocols, in monitoring and reporting adverse events, baseline activity levels and adherence rates to the exercise training.

Our post hoc analyses, however, offer valuable insights into the characteristics of individuals with long COVID who may be more susceptible to adverse events including those PEM-related. Specifically, our findings suggest that those with comorbid cardiac disease and higher PEM scores at baseline may be at greater risk for experiencing adverse events during exercise training and those with comorbid cardiac disease, higher PEM scores at baseline, PEM triggered by mental effort, and a shorter hospital stay may be at greater risk for experiencing PEM-related adverse events during exercise training. To our knowledge, this is the first study to highlight these potential risk factors in this population.

The acceptability of the virtual rehabilitation program was notably high among participants. This was evidenced by the high adherence rate, low dropout rate as well as by the scores and written responses to the satisfaction survey. The convenience, flexibility and accessibility of the virtual format significantly contributed to these adherence rates, allowing participants to integrate the sessions into their schedules and engage with the program more consistently. This type of intervention can be easily implemented across a wide range of settings and has the potential to reach rural, isolated, or under-serviced populations. With appropriate cultural adaptations, it could also be effectively used in different countries.

Our study findings align with previous research indicating that physical rehabilitation interventions lead to improvements in some aspects of physical functioning and in symptoms compared to usual care, and also offer several strengths and unique contributions to the literature. First, although long COVID prevalence among non-hospitalized patients can be as high as 69%,^38^ this cohort had not been included in previous rehabilitation trials comparing exercise training with usual care.^14^ Second, we provided a safe, individualized, symptom-titrated exercise training program and closely monitored for adverse events. Third, all outcome measures in our study align with the Core Outcome Set for long COVID.^39^ Our primary outcome (AM-PAC mobility) is a life impact outcome (recommended in the Core Outcome Set for long COVID) which had not been considered in previous studies. The robust RCT methodology, high adherence rate, and low dropout rate are additional notable study strengths.

Our study has some limitations. Adverse events were not monitored in the control group, and cautious monitoring of adverse events in the intervention group may have resulted in the overreporting of events. Furthermore, the 8-week duration of the program, with sessions three times a week, may not have allowed some participants to fully recover from their fatigue between sessions; indeed, some participants suggested a longer program with more recovery time between sessions in the satisfaction survey. Our program focused on physical rehabilitation and education. Given the comprehensive and multi-dimensional nature of long COVID symptoms,^4^ a multidisciplinary approach might be more effective in addressing common issues such as nutrition, sleep and mental health. Finally, the sample’s high education level and socio-economic status may limit the generalizability of the findings.

Future research should focus on long-term follow-up to assess the sustainability of the observed benefits. Studies exploring the optimal duration of exercise interventions and frequency of sessions to allow for recovery of transient symptom worsening, as well as their integration with other therapeutic approaches, are also warranted. Finally, more research is needed to understand the mechanisms by which exercise impacts long COVID symptoms, which could inform more targeted interventions.

In conclusion, our study did not demonstrate a statistically significant between-group difference in the primary outcome, AM-PAC mobility, however, the proportion of participants achieving the MDC for this measure was twice as high in the intervention group compared to the control group. In addition, our study demonstrated that an 8-week virtual home-based rehabilitation program, which includes long covid-specific education and individualized, symptom-titrated exercise training with cautious monitoring of PEM and adverse events, can improve health status and alleviate common persistent symptoms associated with long COVID such as fatigue, dyspnea, and pain/discomfort. Progression of exercise training is challenging in this population; however, among those who were able to advance during the exercise program, additional improvements were observed in physical functioning and anxiety. The high acceptability and adherence rates indicate that such programs are both feasible and well-received by many individuals with long COVID. Individualized approaches and careful monitoring are essential to accommodate varying levels of symptom severity and avoid symptom exacerbation.

## Data Availability

All data produced in the present study are available upon reasonable request to the authors

## ACKNOWLEDGEMENTS

This study was funded by the Canadian Institutes of Health Research (CIHR)

---

**Table 1 – Supplemental.**
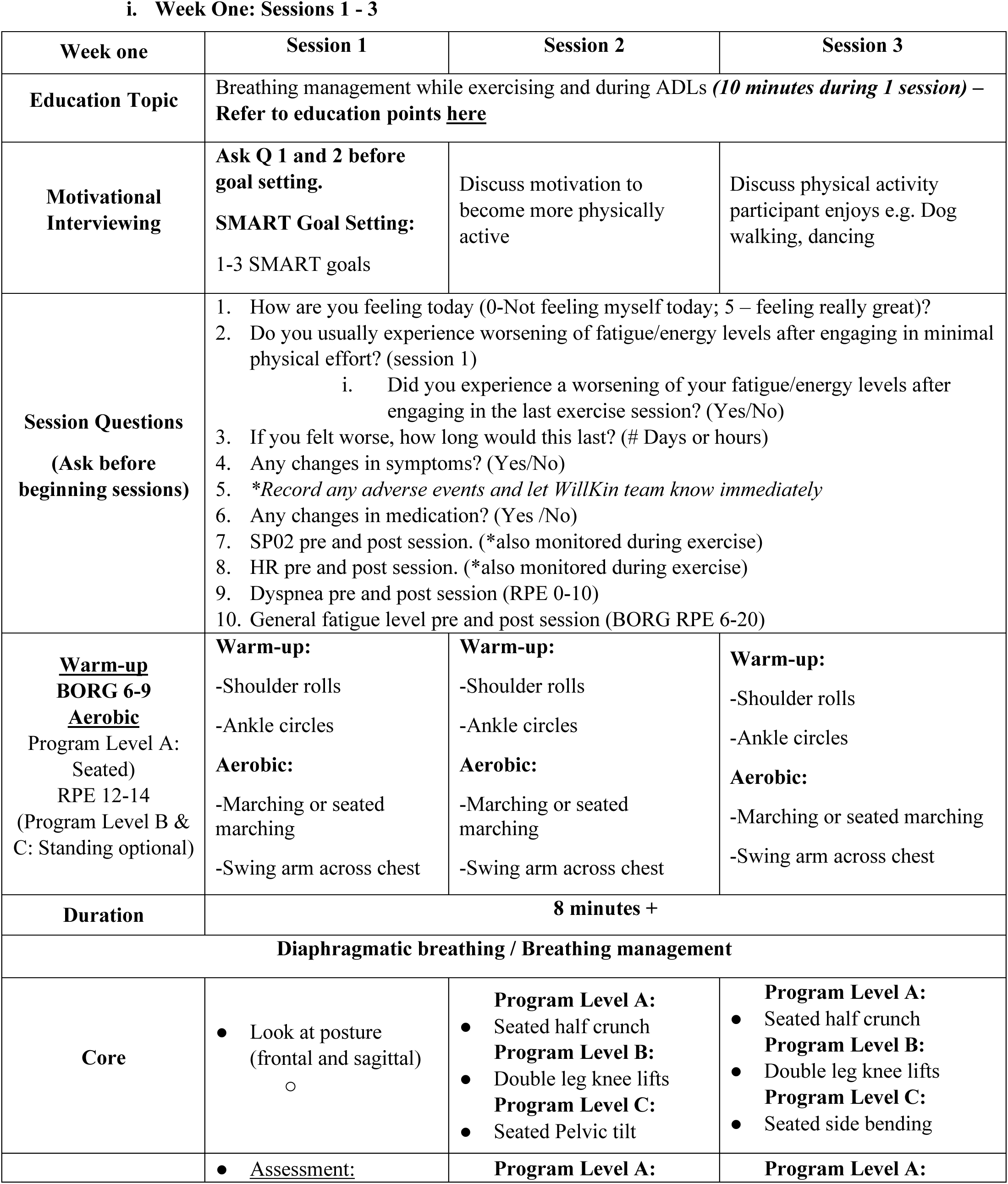
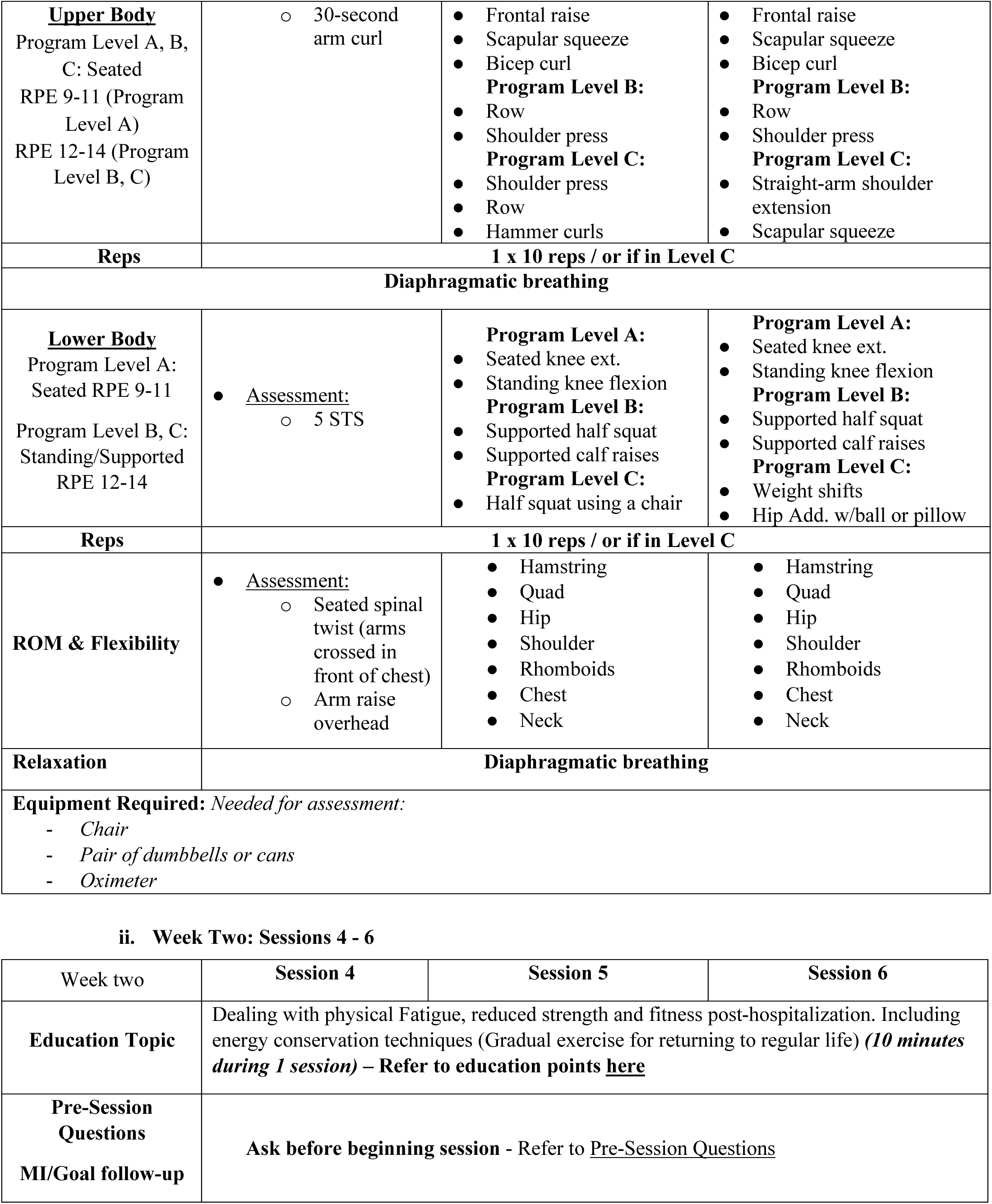
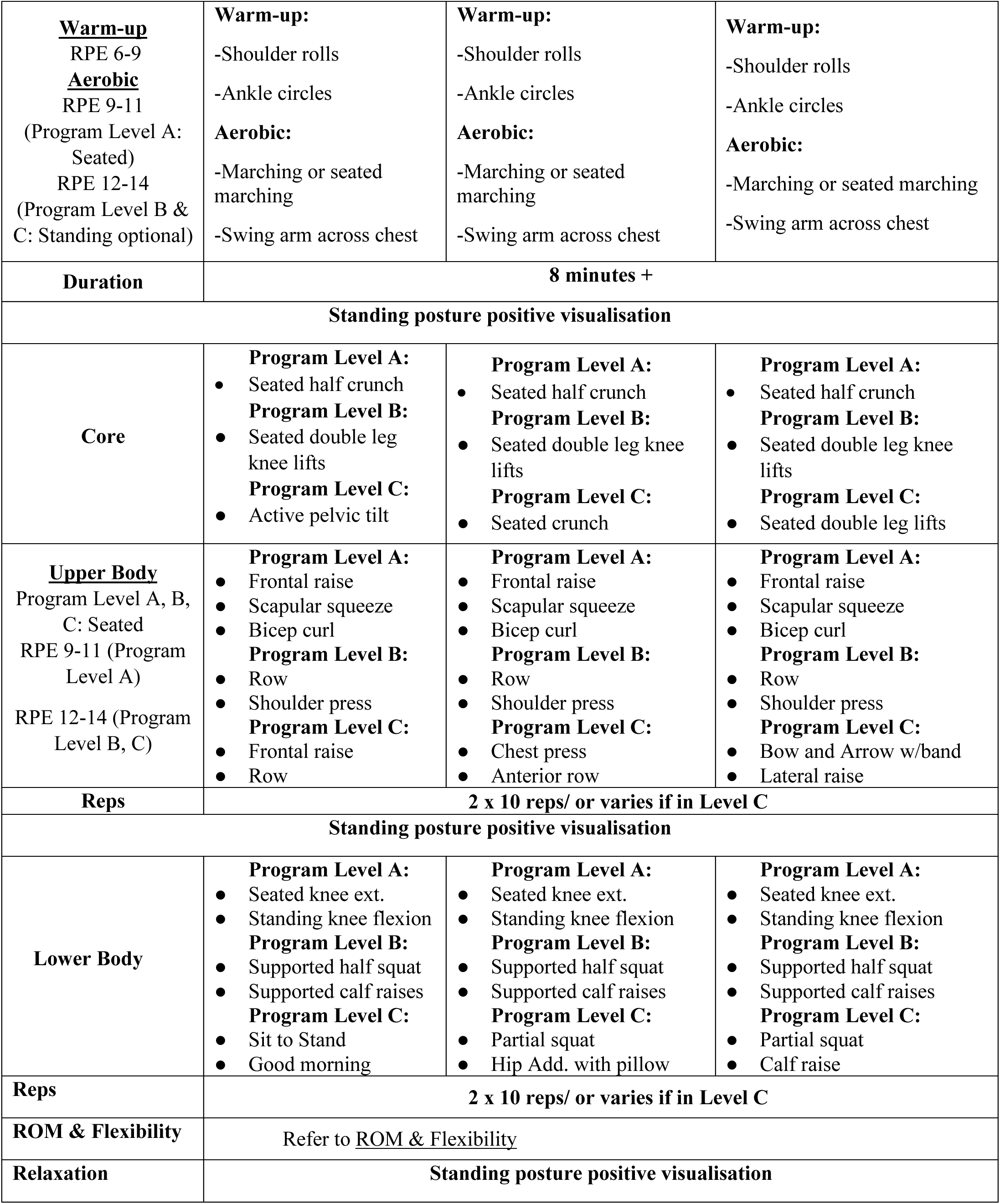
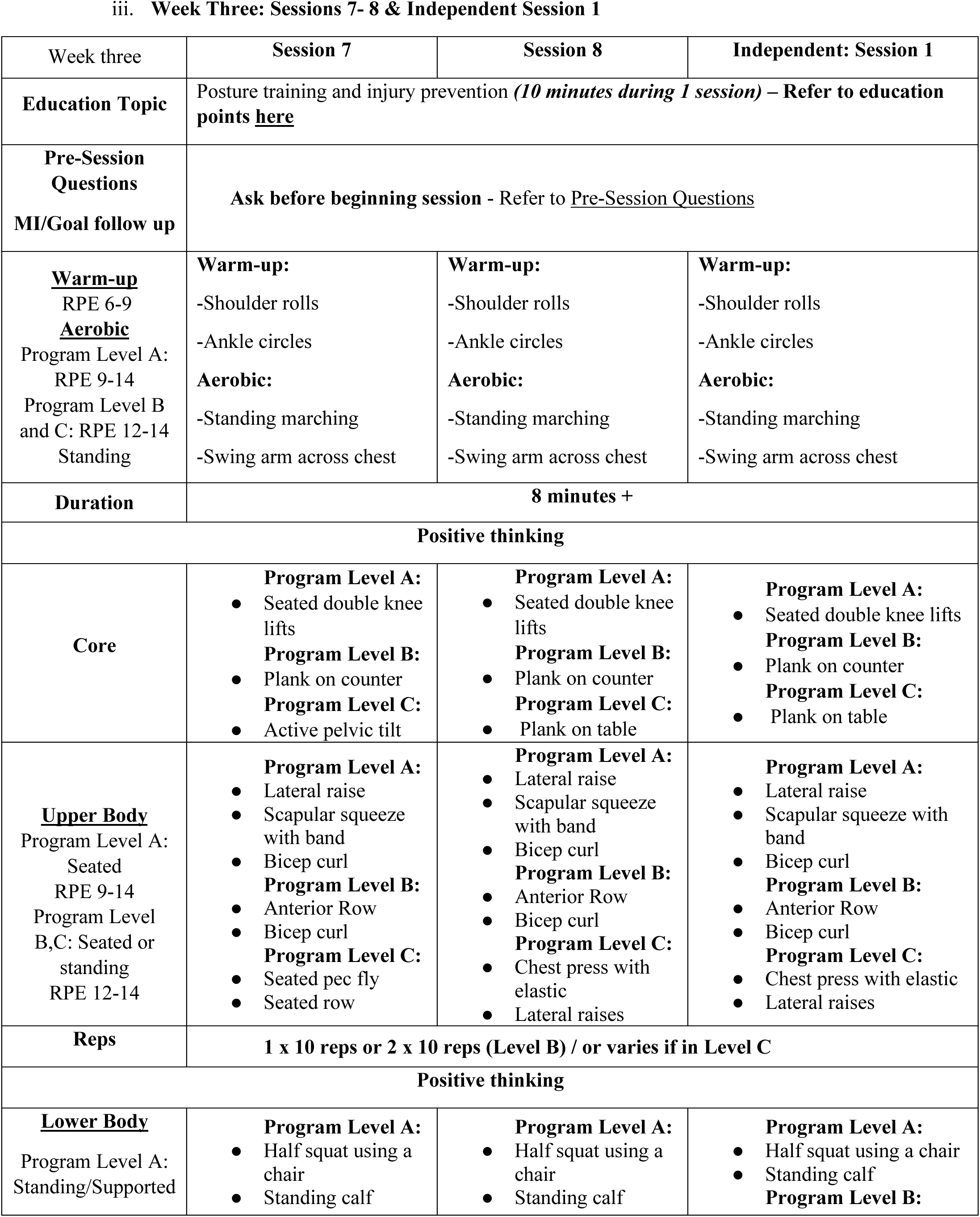
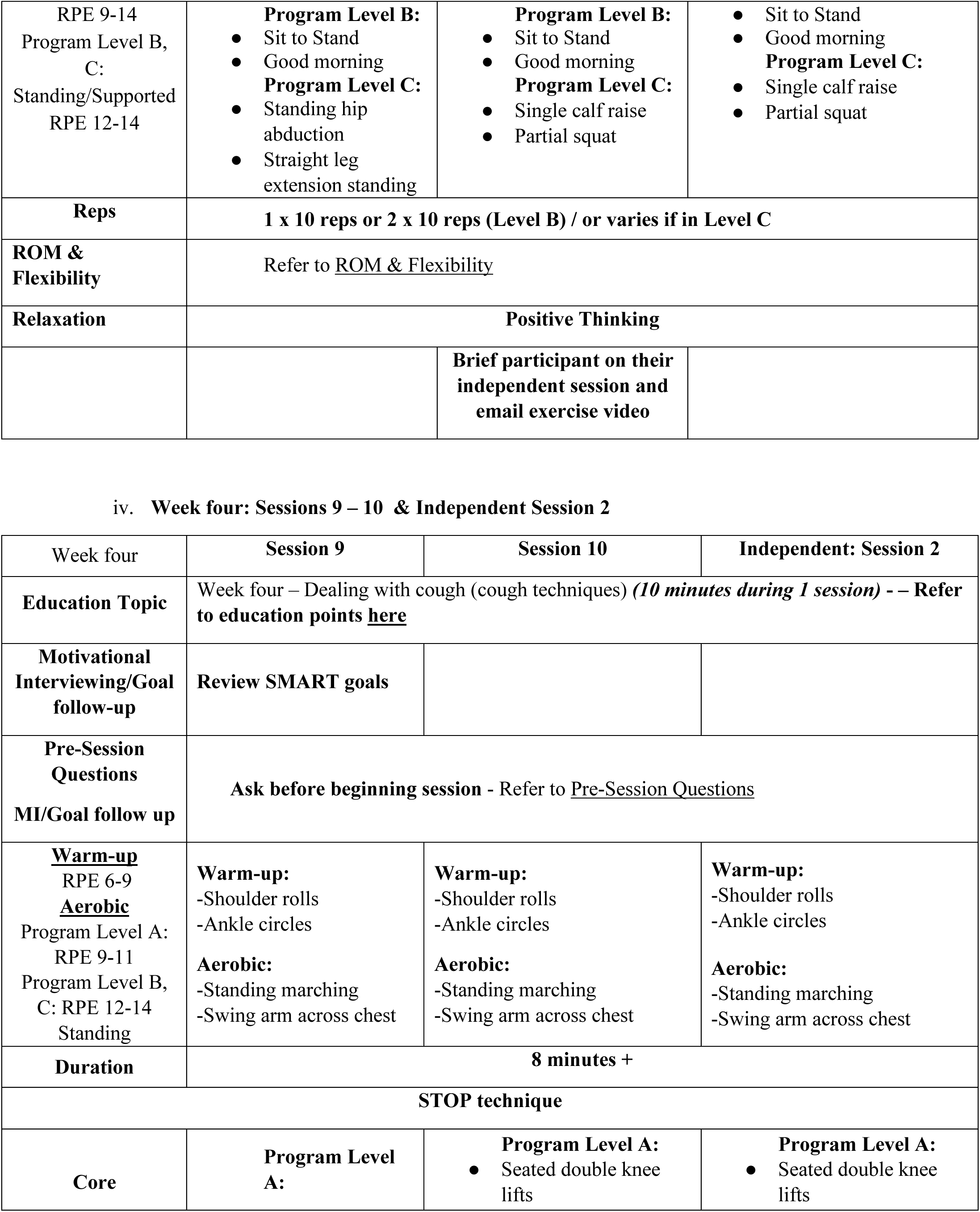
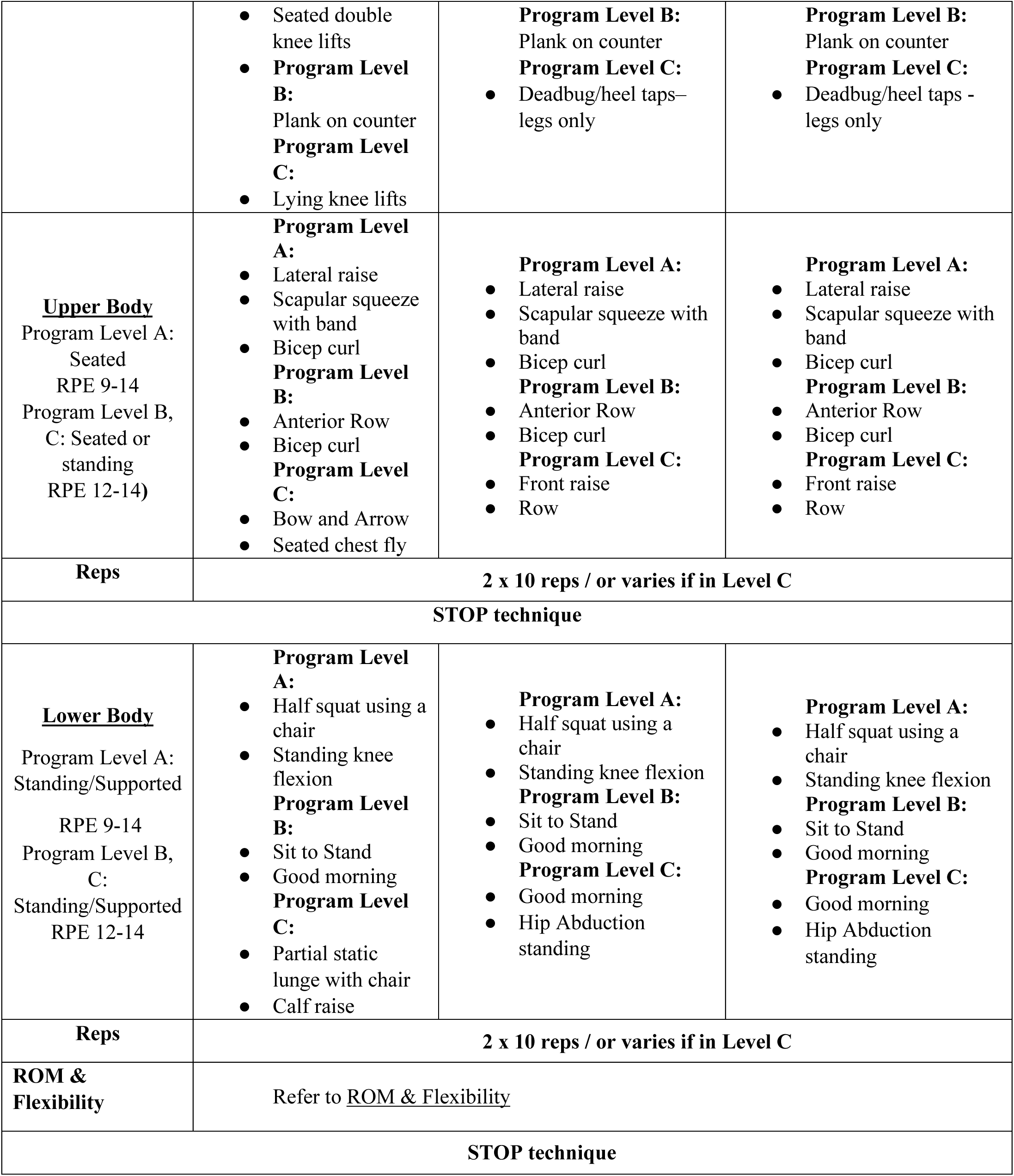
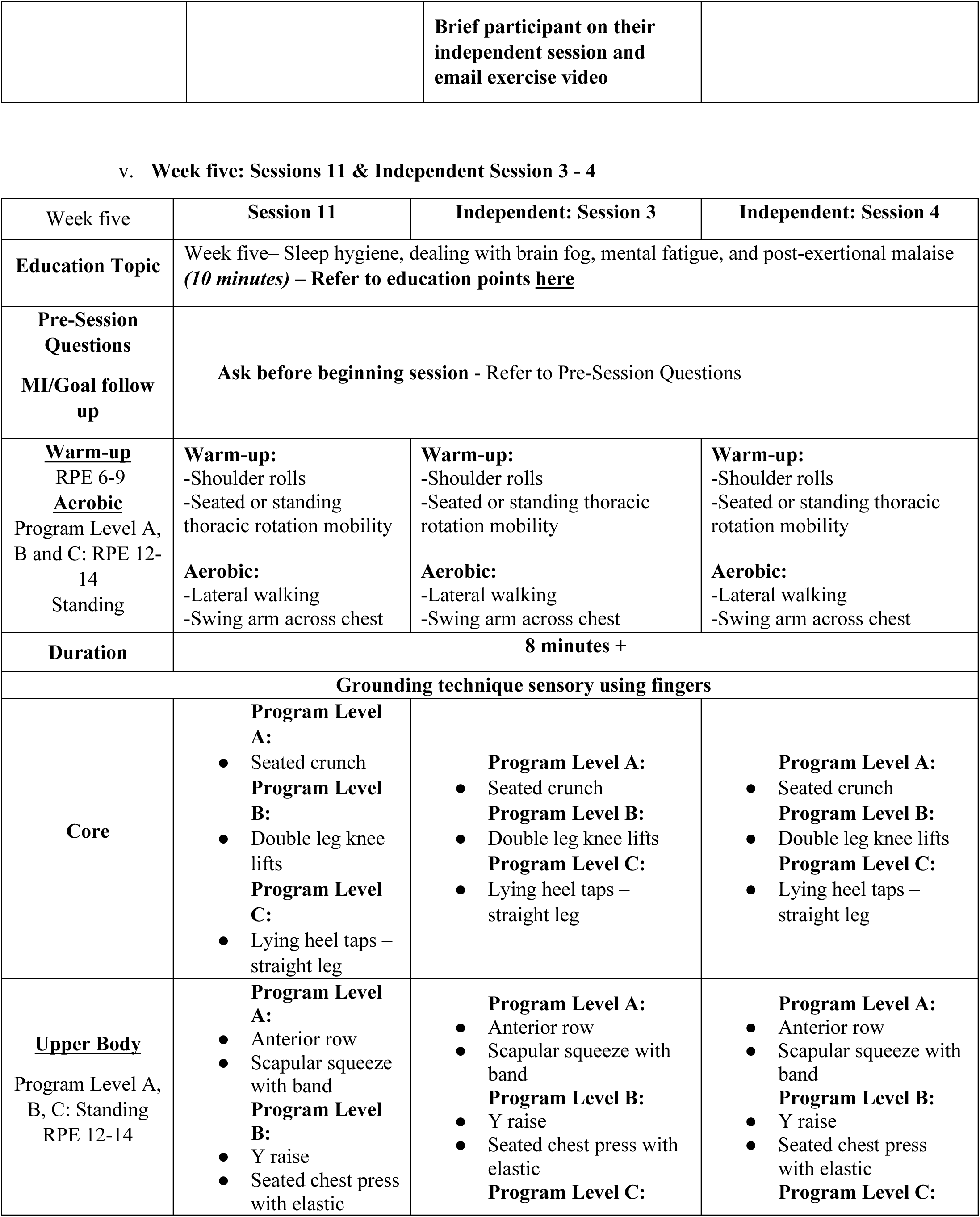
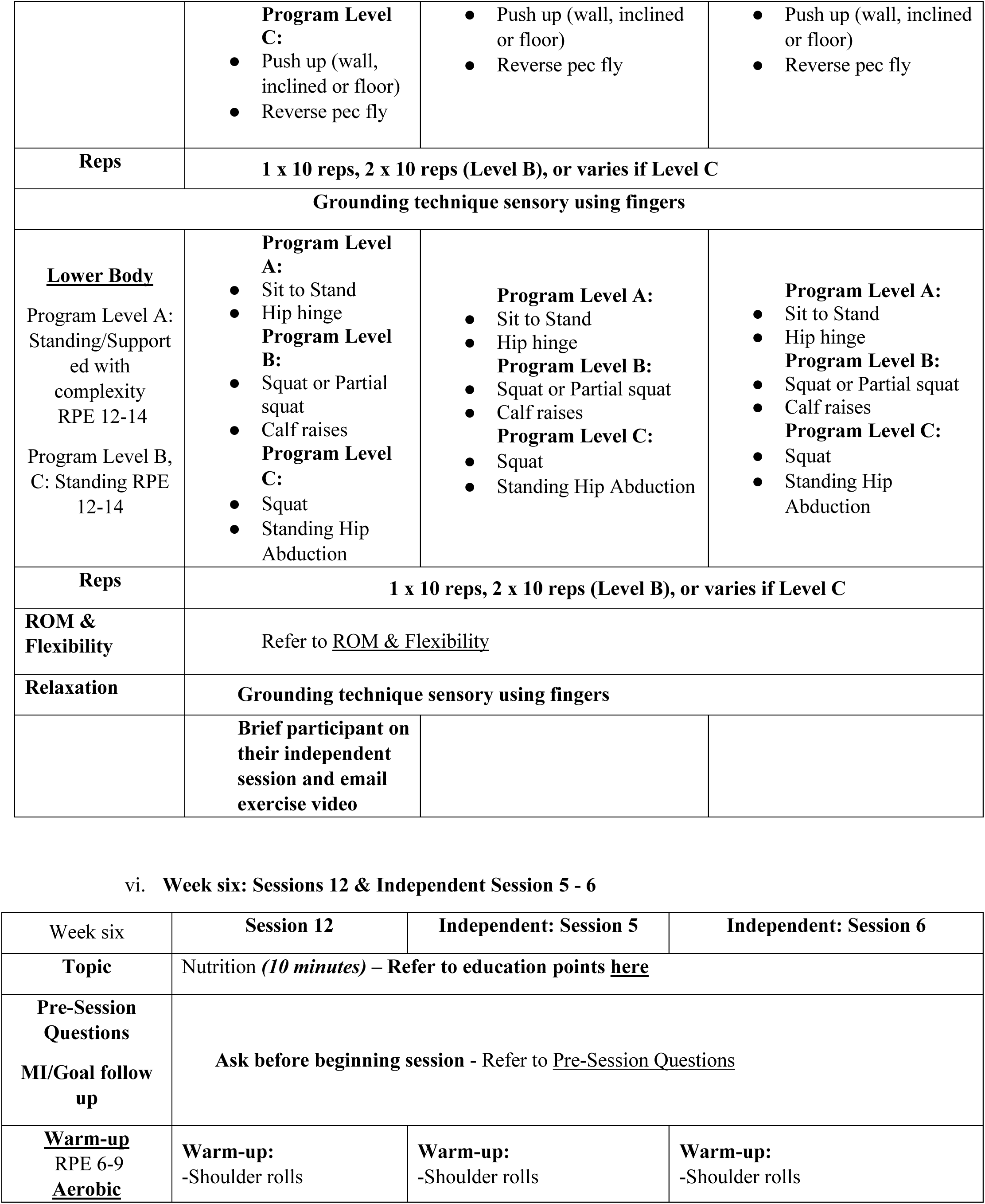
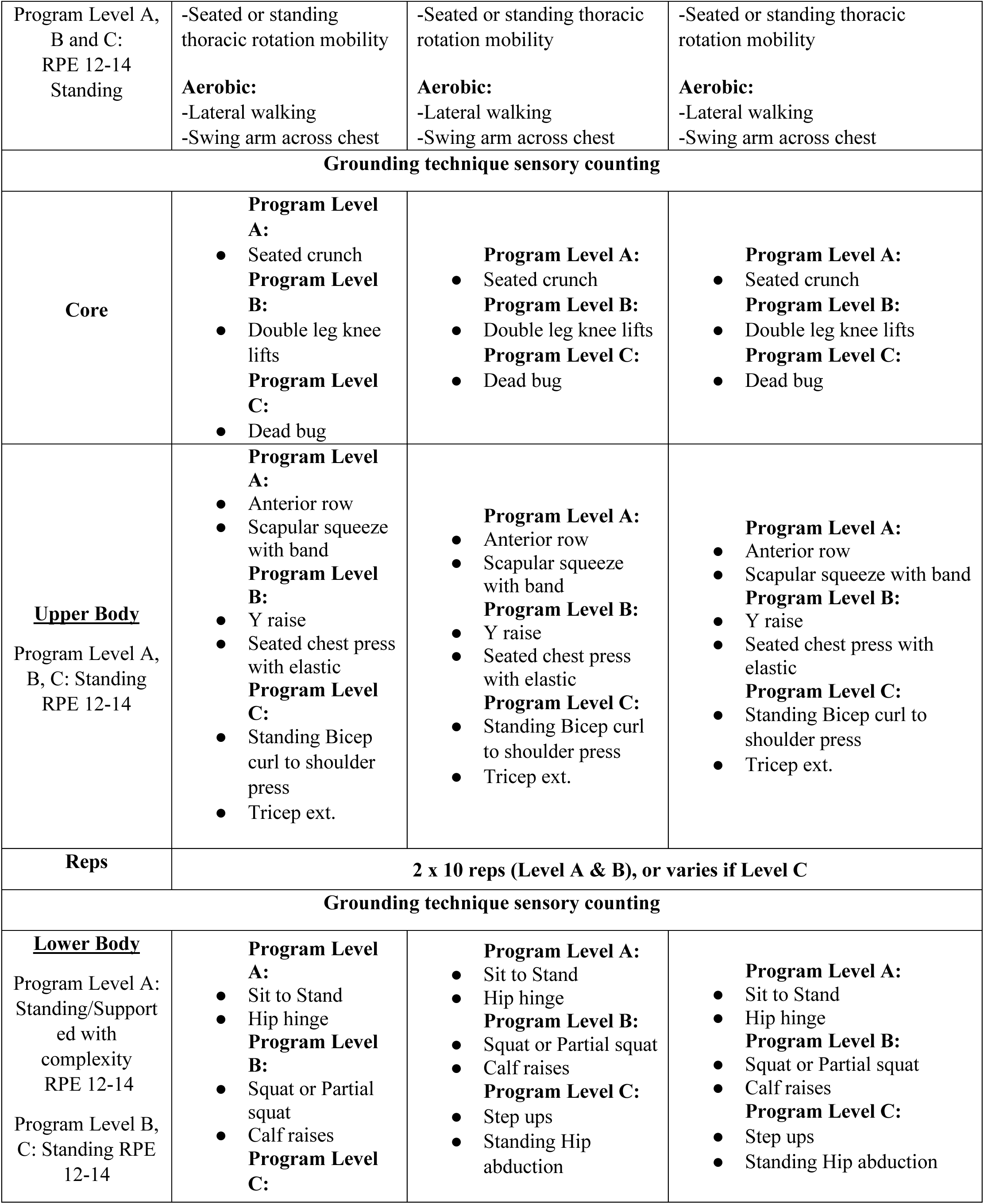
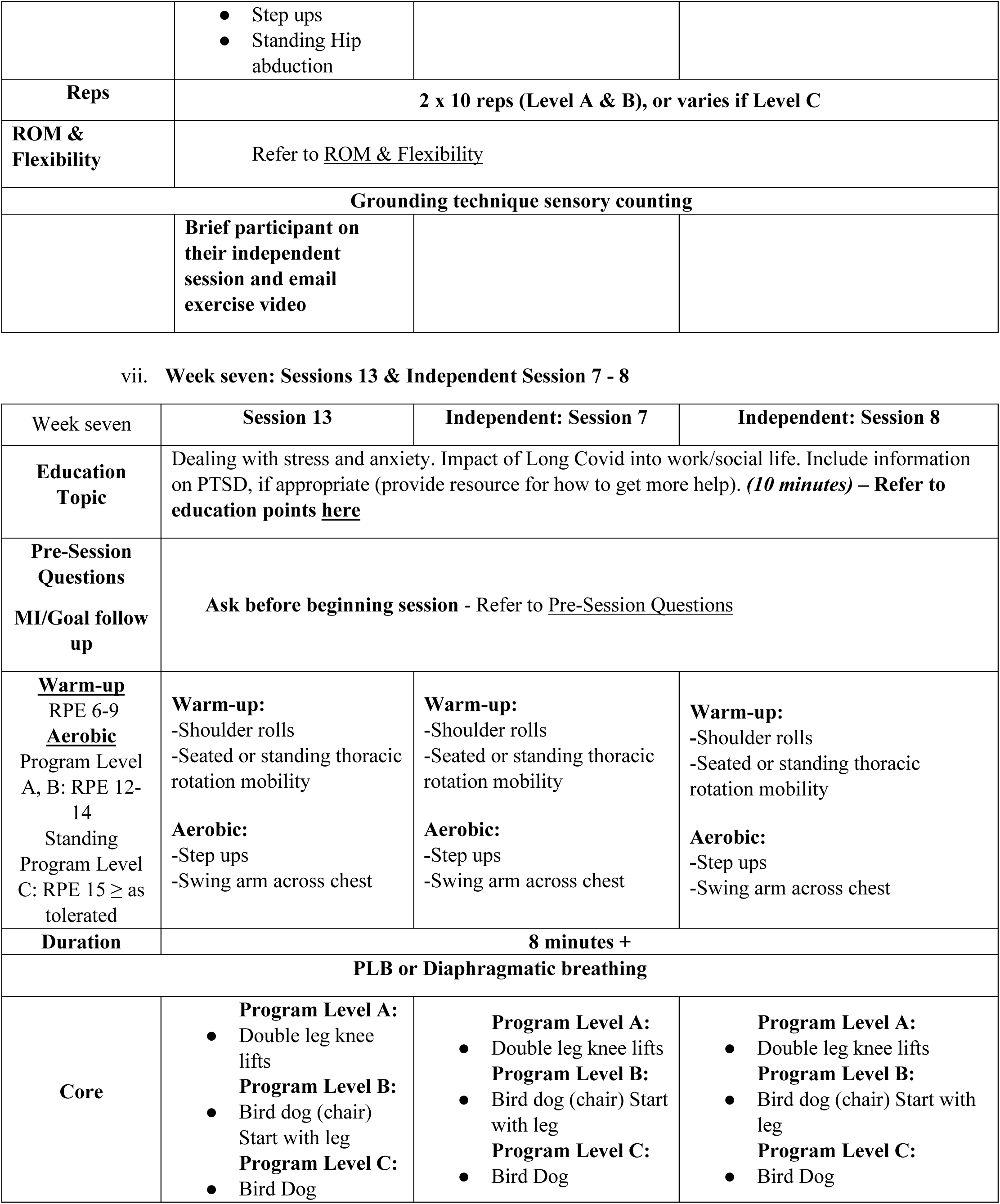
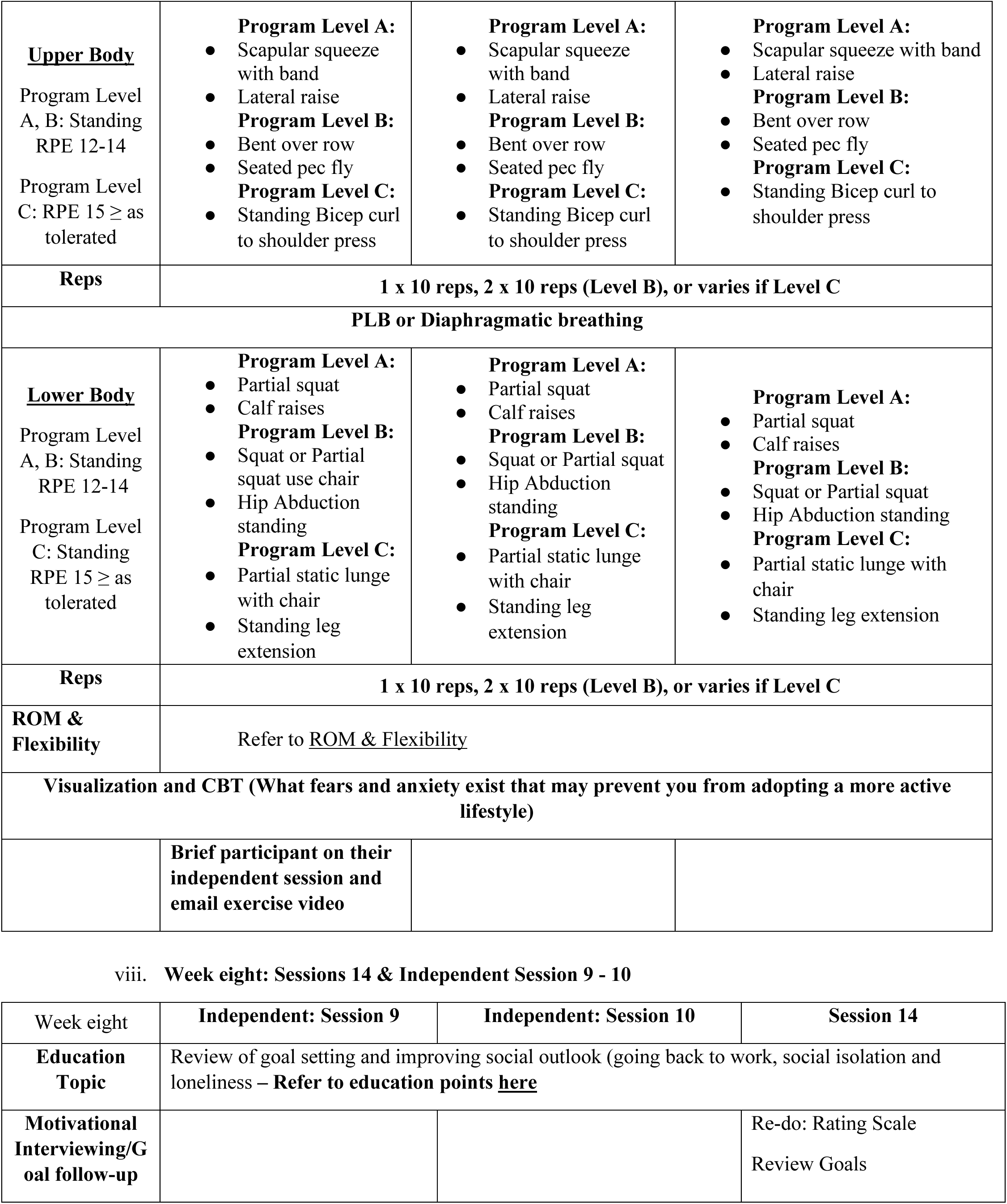
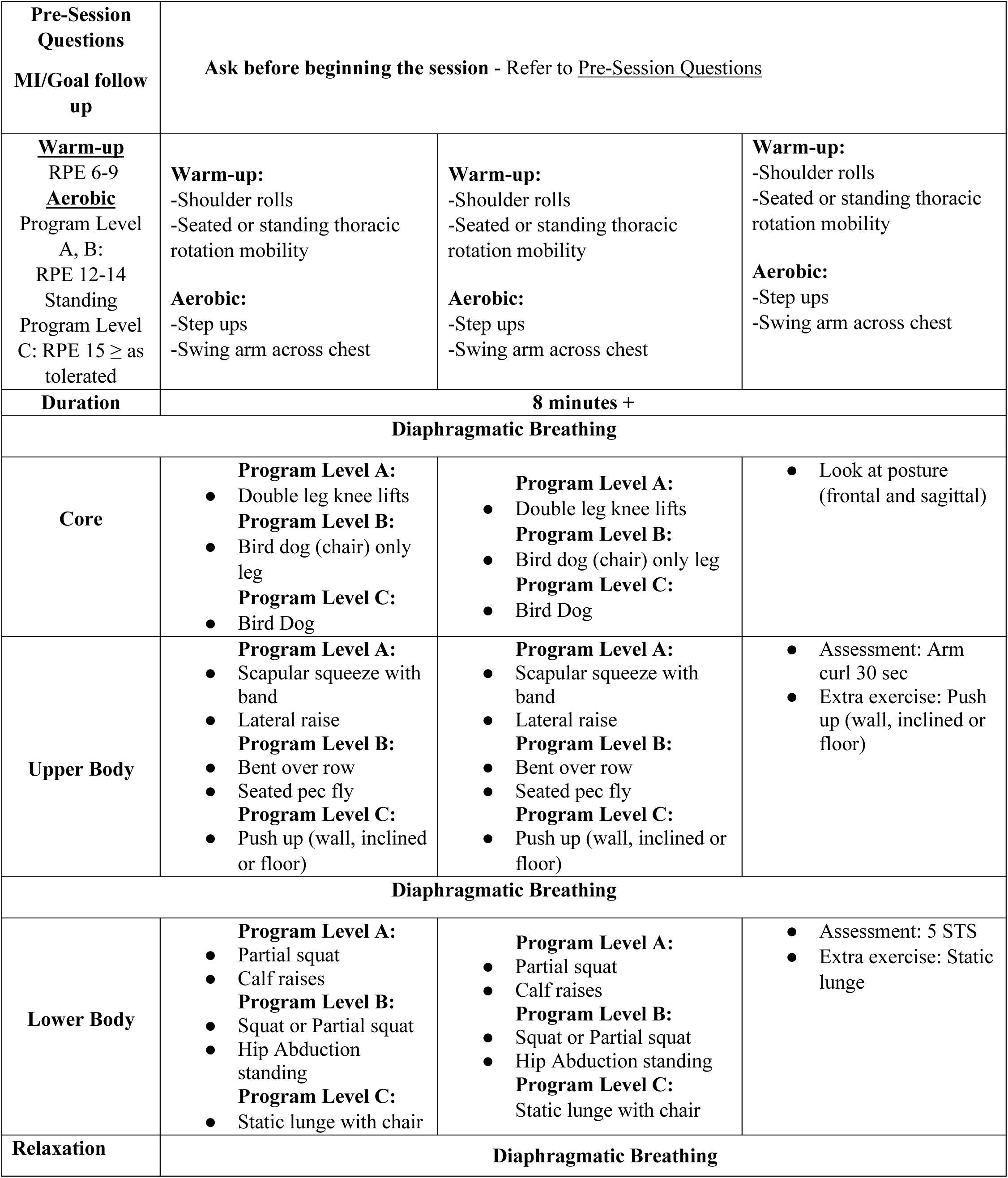
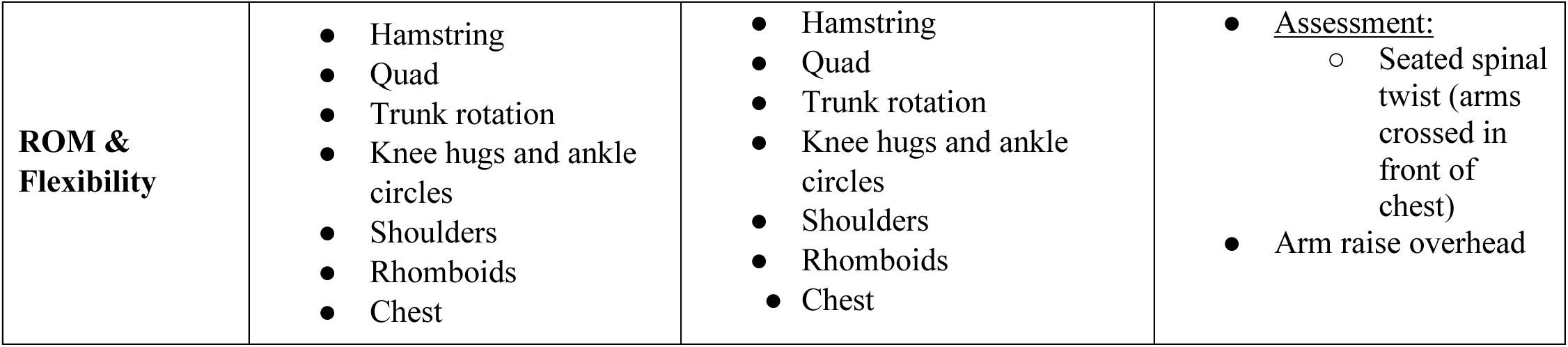
Exercise protocol (Week 1 – 8)

**Table 2 – supplemental.**
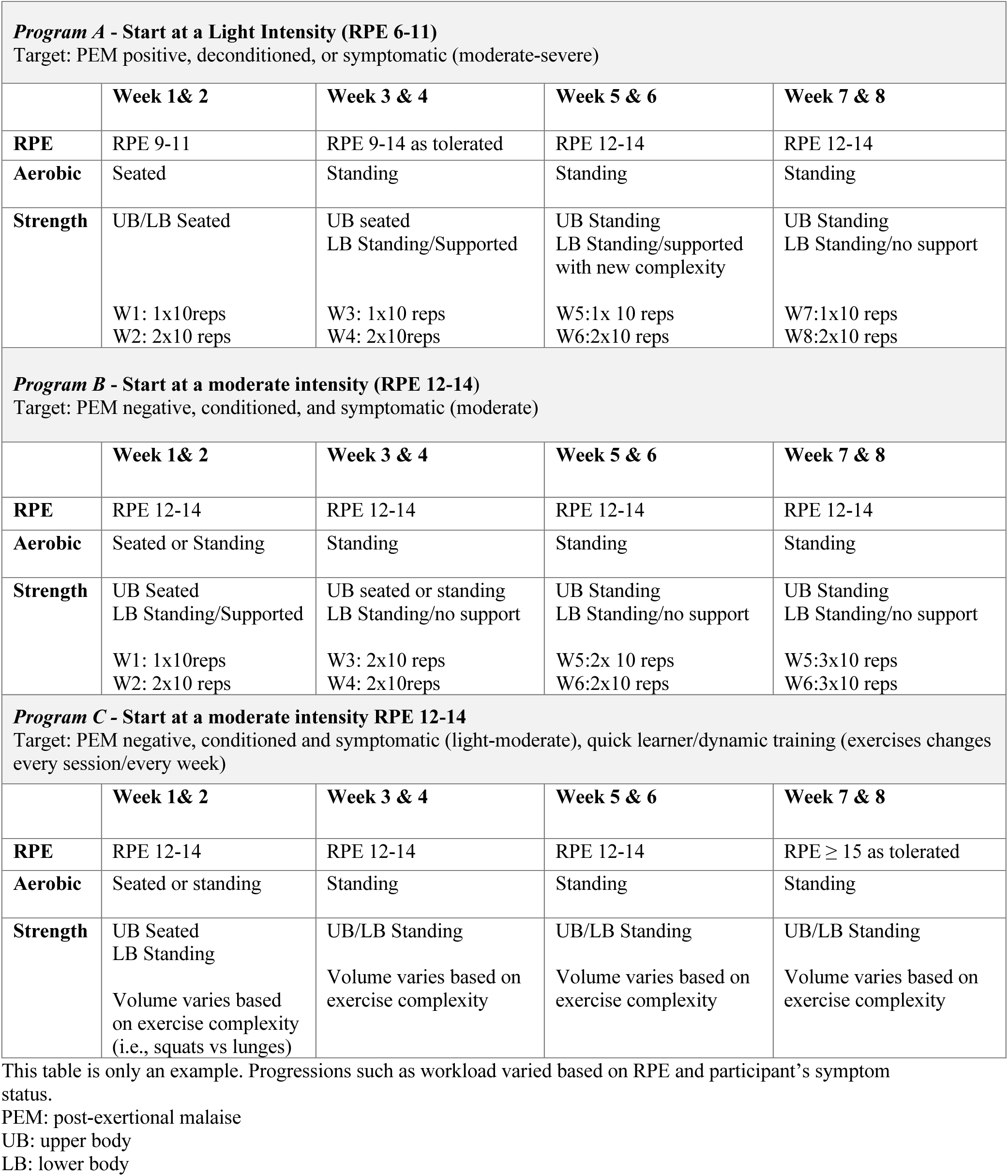
Example of Exercise Progression.

| **Program A - Start at a Light Intensity (RPE 6-11)**<br>Target: PEM positive, deconditioned, or symptomatic (moderate-severe) |  |  |  |  |
| --- | --- | --- | --- | --- |
|  | **Week 1 & 2** | **Week 3 & 4** | **Week 5 & 6** | **Week 7 & 8** |
| **RPE** | RPE 9-11 | RPE 9-14 as tolerated | RPE 12-14 | RPE 12-14 |
| **Aerobic** | Seated | Standing | Standing | Standing |
| **Strength** | UB/LB Seated<br>W1: 1x10reps<br>W2: 2x10 reps | UB seated<br>LB Standing/Supported<br>W3: 1x10 reps<br>W4: 2x10reps | UB Standing<br>LB Standing/supported<br>with new complexity<br>W5:1x 10 reps<br>W6:2x10 reps | UB Standing<br>LB Standing/no support<br>W7:1x10 reps<br>W8:2x10 reps |
| **Program B - Start at a moderate intensity (RPE 12-14)**<br>Target: PEM negative, conditioned, and symptomatic (moderate) |  |  |  |  |
|  | **Week 1 & 2** | **Week 3 & 4** | **Week 5 & 6** | **Week 7 & 8** |
| **RPE** | RPE 12-14 | RPE 12-14 | RPE 12-14 | RPE 12-14 |
| **Aerobic** | Seated or Standing | Standing | Standing | Standing |
| **Strength** | UB Seated<br>LB Standing/Supported<br>W1: 1x10reps<br>W2: 2x10 reps | UB seated or standing<br>LB Standing/no support<br>W3: 2x10 reps<br>W4: 2x10reps | UB Standing<br>LB Standing/no support<br>W5:2x 10 reps<br>W6:2x10 reps | UB Standing<br>LB Standing/no support<br>W5:3x10 reps<br>W6:3x10 reps |
| **Program C - Start at a moderate intensity RPE 12-14**<br>Target: PEM negative, conditioned and symptomatic (light-moderate), quick learner/dynamic training (exercises changes every session/every week) |  |  |  |  |
|  | **Week 1 & 2** | **Week 3 & 4** | **Week 5 & 6** | **Week 7 & 8** |
| **RPE** | RPE 12-14 | RPE 12-14 | RPE 12-14 | RPE ≥ 15 as tolerated |
| **Aerobic** | Seated or standing | Standing | Standing | Standing |
| **Strength** | UB Seated<br>LB Standing<br>Volume varies based<br>on exercise complexity<br>(i.e., squats vs lunges) | UB/LB Standing<br>Volume varies based on<br>exercise complexity | UB/LB Standing<br>Volume varies based on<br>exercise complexity | UB/LB Standing<br>Volume varies based on<br>exercise complexity |

This table is only an example. Progressions such as workload varied based on RPE and participant's symptom status.
PEM: post-exertional malaise
UB: upper body
LB: lower body

**Table 3 – Supplemental.** Adverse events unrelated to the intervention.

| Type* | Not Related |  |  | Total |
| --- | --- | --- | --- | --- |
|  | Grade 1<br>(mild) | Grade 2<br>(moderate) | Grade 3<br>(severe) |  |
| Musculoskeletal | 2 | 1 | 1 | 4 |
| Respiratory | 3 | 7 | 1 | 11 |
| Cardiovascular | 4 | 2 | 0 | 6 |
| Gastrointestinal | 2 | 2 | 0 | 4 |
| Neurological | 0 | 1 | 0 | 1 |
| Psychological | 1 | 1 | 0 | 2 |
| Multisystem† (brain fog, headaches, etc.) | 3 | 4 | 0 | 7 |
| Other (skin issue, unknown) | 1 | 2 | 0 | 3 |

\*adverse events may have been categorized in more than one category
†refers to the multitude of symptoms related to PEM

**Table 4 – Supplemental.** Demographics of those with no AEs vs. those with AEs.

|  | Total in the intervention group (n = 65) | No AEs Group (n = 35) | AEs Group (n = 30) | Mean/proportion difference (95% CI) | Difference for mean±SD/proportion (SE) | P-value |
| --- | --- | --- | --- | --- | --- | --- |
| **Site, n (%)** |  |  |  |  |  |  |
| MUHC | 57 (87.7) | 30 (85.7) | 27 (90.0) | 4.29 (-11.51, 20.09) | 4.29 (8.06) | 0.72 |
| McMaster | 8 (12.3) | 5 (14.3) | 3 (10.0) | -4.29 (-20.09, 11.51) | -4.29 (8.06) | 0.72 |
| **Age (years)** | 48.0 ± 12.0 | 49.2 ± 10.8 | 46.7 ± 13.3 | -2.50 (-8.47, 3.47) | -2.50 ± 12.01 | 0.41 |
| **Sex, n (%)** |  |  |  |  |  |  |
| Female | 46 (70.8) | 25 (71.4) | 21 (70.0) | -1.43 (-23.63, 20.77) | -1.43 (11.33) | 0.90 |
| Male | 19 (29.2) | 10 (28.6) | 9 (30.0) | 1.43 (-20.77, 23.63) | 1.43 (11.33) | 0.90 |
| **Gender, n (%)** |  |  |  |  |  |  |
| Woman | 45 (69.2) | 25 (71.4) | 20 (66.7) | -4.76 (-27.31, 17.79) | -4.76 (11.51) | 0.68 |
| Man | 19 (29.2) | 10 (28.6) | 9 (30.0) | 1.43 (-20.77, 23.63) | 1.43 (11.33) | 0.90 |
| Gender queer/Gender non-conforming | 1 (1.5) | 0 (0.0) | 1 (3.3) | 3.33 (-3.09, 9.76) | 3.33 (3.28) | 0.46 |
| **Ethnicity, n (%)** |  |  |  |  |  |  |
| Asian | 3 (4.8) | 2 (5.9) | 1 (3.6) | -2.31 (-12.79, 8.17) | -2.31 (5.35) | 1.00 |
| Black | 0 (0.0) | 0 (0.0) | 0 (0.0) | - | - | - |
| Caucasian | 55 (88.7) | 30 (88.2) | 25 (89.3) | 1.05 (-14.71, 16.82) | 1.05 (8.04) | 1.00 |
| Latino American | 0 (0.0) | 0 (0.0) | 0 (0.0) | - | - | - |
| Middle East/North African | 3 (4.8) | 1 (2.9) | 2 (7.1) | 4.20 (-6.90, 15.30) | 4.20 (5.66) | 0.58 |
| Other | 1 (1.6) | 1 (2.9) | 0 (0.0) | -2.94 (-8.62, 2.74) | -2.94 (2.9) | 1.00 |
| **BMI, kg/m<sup>2</sup>** | 29.2 ± 9.2 | 30.0 ± 10.4 | 28.2 ± 7.5 | -1.85 (-6.42, 2.72) | -1.85 ± 9.19 | 0.65 |
| **Education Highest degree attained, n (%)** |  |  |  |  |  |  |
| Graduate | 17 (26.2) | 10 (28.6) | 7 (23.3) | -5.24 (-26.52, 16.05) | -5.24 (10.86) | 0.63 |
| Undergraduate | 22 (33.8) | 12 (34.3) | 10 (33.3) | -0.95 (-24.01, 22.11) | -0.95 (11.77) | 0.94 |
| College | 15 (23.1) | 10 (28.6) | 5 (16.7) | -11.90 (-31.95, 8.14) | -11.90 (10.23) | 0.26 |
| High school | 9 (13.8) | 2 (5.7) | 7 (23.3) | 17.62 (-0.14, 17.76) | 17.62 (8.66) | 0.07 |
| Other | 2 (3.1) | 1 (2.9) | 1 (3.3) | 0.48 (-7.99, 8.95) | 0.48 (4.32) | 1.00 |
| **Household income, n (%)** |  |  |  |  |  |  |
| Less than \$25,000 | 2 (3.8) | 1 (3.6) | 1 (4.0) | 0.43 (-9.88, 10.74) | 0.43 (5.26) | 1.00 |
| \$25,000 to \$50,000 | 7 (13.2) | 3 (10.7) | 4 (16.0) | 5.29 (-13.09, 23.66) | 5.29 (9.38) | 0.69 |
| \$50,001 to \$75,000 | 14 (26.4) | 5 (17.9) | 9 (36.0) | 18.14 (-5.42, 41.71) | 18.14 (12.02) | 0.13 |
| \$75,001 to \$100,000 | 11 (20·8) | 6 (21·4) | 5 (20·0) | -1·43 (-23·27, 20·41) | -1·43 (11·14) | 0·90 |
| \$100,001 or more | 19 (35·8) | 13 (46·4) | 6 (24·0) | -22·43 (-47·36, 2·50) | -22·43 (12·72) | 0·09 |
| **Work status, n (%)** |  |  |  |  |  |  |
| Employed/self-employed | 44 (67·7) | 26 (74·3) | 18 (60·0) | -14·29 (-37·02, 8·45) | -14·29 (11·60) | 0·22 |
| Employed/self-employed yet not working | 37 (84·1) | 21 (80·8) | 16 (88·9) | 8·12 (-12·86, 29·10) | 8·12 (10·71) | 0·68 |
| Unemployed | 6 (9·2) | 1 (2·9) | 5 (16·7) | 13·81 (-0·62, 28·24) | 13·81 (7·36) | 0·09 |
| Studying | 1 (1·5) | 0 (0·0) | 1 (3·3) | 3·33 (-30·41, 37·08) | 3·33 (3·28) | 0·46 |
| Retired | 8 (12·3) | 6 (17·1) | 2 (6·7) | -10·48 (-25·82, 4·87) | -10·48 (7·83) | 0·27 |
| Other | 6 (9·2) | 2 (5·7) | 4 (13·3) | 7·62 (-6·77, 22·01) | 7·62 (7·34) | 0·40 |
| **Cigarette smoking status, n (%)** |  |  |  |  |  |  |
| Current smoker | 0 (0·0) | 0 (0·0) | 0 (0·0) | - | - | - |
| Previous smoker | 25 (38·5) | 11 (31·4) | 14 (46·7) | 15·24 (-8·33, 38·80) | 15·24 (12·02) | 0·21 |
| Never smoked | 40 (61·5) | 24 (68·6) | 16 (53·3) | -15·24 (-38·80, 8·33) | -15·24 (12·02) | 0·21 |
| **Comorbidities, n (%)** |  |  |  |  |  |  |
| Hypertension | 12 (18·5) | 8 (22·9) | 4 (13·3) | -9·52 (-28·00, 8·96) | -9·52 (9·43) | 0·36 |
| Diabetes | 4 (6·2) | 2 (5·7) | 2 (6·7) | 0·95 (-10·83, 12·73) | 0·95 (6·01) | 1·00 |
| Musculoskeletal disease | 13 (20·0) | 5 (14·3) | 8 (26·7) | 12·38 (-7·24, 32·00) | 12·38 (10·01) | 0·21 |
| Cardiac disease | 4 (6·2) | 0 (0·0) | 4 (13·3) | 13·33 (11·70, 25·50) | 13·33 (6·21) | 0·04* |
| Cancer | 0 (0·0) | 0 (0·0) | 0 (0·0) | - | - | - |
| Respiratory disease (other than COVID) | 11 (16·9) | 5 (14·3) | 6 (20·0) | 5·71 (-12·71, 24·13) | 5·71 (9·40) | 0·54 |
| Number of comorbidities, median (Q1, Q3) | 0·0 (0·0, 1·0) | 0·0 (0·0, 2·0) | 1·0 (0·0, 1·0) | 0·03 (-0·64, 0·71) | 0·03 ± 1·36 | 0·67 |
| Number of comorbidities as category |  |  |  |  |  |  |
| 0 | 33 (50·8) | 19 (54·3) | 14 (46·7) | -7·62 (-31·93, 16·69) | -7·62 (12·40) | 0·54 |
| 1 | 16 (24·6) | 7 (20·0) | 9 (30·0) | 10·00 (-11·08, 31·08) | 10·00 (10·76) | 0·35 |
| 2+ | 16 (24·6) | 9 (25·7) | 7 (23·3) | -2·38 (-23·33, 18·56) | -2·38 (10·69) | 0·82 |
| **Taking prescribed medication, n (%)** | 55 (84·6) | 28 (80·0) | 27 (90·0) | 10·00 (-7·05, 27·05) | 10·00 (8·70) | 0·32 |
| **COVID Frequency, median (Q1, Q3)** | 1·0 (1·0, 2·0) | 1·0 (1·0, 2·0) | 1·0 (1·0, 2·0) | -0·04 (-0·37, 0·29) | -0·04 ± 0·64 | 0·67 |
| **Long covid duration, in months, median (Q1, Q3)** | 13·6 (7·8, 21·1) | 14·8 (6·6, 23·6) | 12·9 (8·4, 20·8) | -1·59 (-6·14, 2·97) | -1·59 ± 9·08 | 0·71 |
| **Number of symptoms, median (Q1, Q3)** | 5·0 (3·0, 7·0) | 4·0 (2·0, 7·0) | 5·0 (4·0, 7·0) | 0·81 (-1·16, 2·79) | 0·81 ± 3·96 | 0·12 |
| **Hospitalization** |  |  |  |  |  |  |
| Hospitalized due to COVID, n (%) |  |  |  |  |  |  |
| 0 | 51 (78·5) | 26 (74·3) | 25 (83·3) | 9·05 (-10·64, 28·73) | 9·05 (10·04) | 0·38 |
| 1 | 11 (16·9) | 7 (20·0) | 4 (13·3) | -6·67 (-24·65, 11·32) | -6·67 (9·18) | 0·53 |
| 2+ | 3 (4·6) | 2 (5·7) | 1 (3·3) | -2·38 (-12·40, 7·64) | -2·38 (5·11) | 1·00 |
| 1st Hospitalization LOS | 4·0 ± 16·2 | 5·8 ± 20·9 | 1·9 ± 7·5 | -3·84 (-11·88, 4·20) | -3·84 ± 16·17 | 0·22 |
| ≥1 ICU admission for COVID, n (%) | 6 (9·2) | 4 (11·4) | 2 (6·7) | -4·76 (-18·57, 9·05) | -4·76 (7·05) | 0·68 |
| 1st ICU LOS | 1·9 ± 7·4 | 2·1 ± 7·5 | 1·5 ± 7·3 | -0·61 (-4·32, 3·10) | -0·61 ± 7·46 | 0·52 |
| **Number of vaccinations, median (Q1, Q3)** | 3·0 (2·0, 4·0) | 4·0 (3·0, 4·0) | 3·0 (2·0, 4·0) | -0·26 (-0·93, 0·42) | -0·26 ± 1·21 | 0·55 |
| **Supplemental oxygen (O<sub>2</sub>), n (%)** | 0 (0·0) | 0 (0·0) | 0 (0·0) | - | - | - |
| **Current level of activity, n (%)** |  |  |  |  |  |  |
| Vigorously active | 0 (0·0) | 0 (0·0) | 0 (0·0) | - | - | - |
| Moderately active | 15 (23·1) | 9 (25·7) | 6 (20·0) | -5·71 (-26·07, 14·65) | -5·71 (10·39) | 0·59 |
| Seldom active | 50 (76·9) | 26 (74·3) | 24 (80·0) | 5·71 (-14·65, 26·07) | 5·71 (10·39) | 0·59 |
| **BDI Total score (0 - 12), median (Q1, Q3)** | 4·0 (2·0, 5·0) | 4·0 (2·0, 6·0) | 3·5 (2·0, 4·0) | -0·82 (-2·06, 0·42) | -0·82 ± 2·49 | 0·25 |
| **DSQ-PEM** |  |  |  |  |  |  |
| PEM (sum of all scores) (0 - 5), median (Q1, Q3) | 4·0 (3·0, 5·0) | 4·0 (2·0, 5·0) | 5·0 (3·0, 5·0) | 1·01 (0·26, 1·76) | 1·01 ± 1·51 | 0·04* |
| PEM score, n (%) |  |  |  |  |  |  |
| 0 | 2 (3·1) | 0 (0·0) | 2 (6·7) | 6·67 (-2·26, 15·59) | 6·67 (4·55) | 0·21 |
| 1-2 | 13 (20·0) | 10 (28·6) | 3 (10·0) | -18·57 (-36·99, 0·15) | -18·57 (9·40) | 0·07 |
| 3-5 | 50 (76·9) | 25 (71·4) | 25 (83·3) | 11·90 (-8·14, 31·95) | 11·90 (10·23) | 0·26 |
| PEM present with minimal physical efforts, n (%) | 59 (90·8) | 31 (88·6) | 28 (93·3) | 4·76 (-9·05, 18·57) | 4·76 (7·05) | 0·68 |
| PEM present with mental effort, n (%) | 55 (84·6) | 27 (77·1) | 28 (93·3) | 16·19 (-0·34, 32·72) | 16·19 (8·43) | 0·09 |

\*p-value < 0·05
Data are presented as mean ± SD unless otherwise specified
AE: Adverse Event; MUHC: McGill University Health Centre; BMI: body mass index; LOS: length of stay; ICU: intensive care unit; BDI: Beck Depression Inventory; DSQ-PEM: DePaul Symptom Questionnaire - post-exertional malaise.

**Table 5 – Supplemental.** Demographics of Those with No Adverse Events vs. Those with PEM-Related Adverse Events.

|  | Total in the intervention group (n = 57) | No AEs Group (n = 35) | PEM AEs Group (n = 22) | Mean/proportion difference (95% CI) | Difference for mean±SD/proportion (SE) | P-value |
| --- | --- | --- | --- | --- | --- | --- |
| **Site, n (%)** |  |  |  |  |  |  |
| MUHC | 51 (89·5) | 30 (85·7) | 21 (95·5) | 9·74 (-4·76, 24·24) | 9·74 (7·40) | 0·39 |
| McMaster | 6 (10·5) | 5 (14·3) | 1 (4·5) | -9·74 (-24·24, 4·76) | -9·74 (7·40) | 0·39 |
| **Age (years)** | 47·8 ± 10·9 | 49·2 ± 10·8 | 45·6 ± 11·0 | -3·56 (-9·50, 2·37) | -3·56 ± 10·88 | 0·23 |
| **Sex, n (%)** |  |  |  |  |  |  |
| Female | 42 (73·7) | 25 (71·4) | 17 (77·3) | 5·84 (-17·19, 28·88) | 5·84 (11·75) | 0·63 |
| Male | 15 (26·3) | 10 (28·6) | 5 (22·7) | -5·84 (-28·88, 17·19) | -5·84 (11·75) | 0·63 |
| **Gender, n (%)** |  |  |  |  |  |  |
| Woman | 42 (73·7) | 25 (71·4) | 17 (77·3) | 5·84 (-17·19, 28·88) | 5·84 (11·75) | 0·63 |
| Man | 15 (26·3) | 10 (28·6) | 5 (22·7) | -5·84 (-28·88, 17·19) | -5·84 (11·75) | 0·63 |
| Genderqueer/Gender non-conforming | 0 (0·0) | 0 (0·0) | 0 (0·0) | - | - | - |
| **Ethnicity, n (%)** |  |  |  |  |  |  |
| Asian | 3 (5·6) | 2 (5·9) | 1 (5·0) | -0·88 (-13·28, 11·52) | -0·88 (6·33) | 1·00 |
| Black | 0 (0·0) | 0 (0·0) | 0 (0·0) | - | - | - |
| Caucasian | 47 (87·0) | 30 (88·2) | 17 (85·0) | -3·24 (-22·27, 15·80) | -3·24 (9·71) | 1·00 |
| Latino American | 0 (0·0) | 0 (0·0) | 0 (0·0) | - | - | - |
| Middle East/North African | 3 (5·6) | 1 (2·9) | 2 (10·0) | 7·06 (-7·26, 21·38) | 7·06 (7·31) | 0·55 |
| Other | 1 (1·9) | 1 (2·9) | 0 (0·0) | -2·94 (-8·62, 2·74) | -2·94 (2·9) | 1·00 |
| **BMI, kg/m<sup>2</sup>** | 28·8 ± 9·2 | 30·0 ± 10·4 | 27·0 ± 6·7 | -3·05 (-8·04, 1·94) | -3·05 ± 9·16 | 0·37 |
| **Education Highest degree attained, n (%)** |  |  |  |  |  |  |
| Graduate | 15 (26·3) | 10 (28·6) | 5 (22·7) | -5·84 (-28·88, 17·19) | -5·84 (11·75) | 0·63 |
| Undergraduate | 22 (38·6) | 12 (34·3) | 10 (45·5) | 11·17 (-14·91, 37·25) | 11·17 (13·31) | 0·40 |
| College | 14 (24·6) | 10 (28·6) | 4 (18·2) | -10·39 (-32·38, 11·60) | -10·39 (11·22) | 0·53 |
| High school | 5 (8·8) | 2 (5·7) | 3 (13·6) | 7·92 (-8·35, 24·19) | 7·92 (8·30) | 0·36 |
| Other | 1 (1·8) | 1 (2·9) | 0 (0·0) | -2·86 (-8·38, 2·66) | -2·86 (2·82) | 1·00 |
| **Household income, n (%)** |  |  |  |  |  |  |
| Less than \$25,000 | 2 (4·2) | 1 (3·6) | 1 (5·0) | 1·43 (-10·34, 13·20) | 1·43 (6·00) | 1·00 |
| \$25,000 to \$50,000 | 4 (8·3) | 3 (10·7) | 1 (5·0) | -5·71 (-20·63, 9·20) | -5·71 (7·61) | 0·63 |
| \$50,001 to \$75,000 | 13 (27·1) | 5 (17·9) | 8 (40·0) | 22·14 (-3·59, 47·88) | 22·14 (13·13) | 0·11 |
| \$75,001 to \$100,000 | 11 (22·9) | 6 (21·4) | 5 (25·0) | 3·57 (-20·74, 27·88) | 3·57 (12·40) | 1·00 |
| \$100,001 or more | 18 (37·5) | 13 (46·4) | 5 (25·0) | -21·43 (-47·91, 5·05) | -21·43 (13·51) | 0·23 |
| **Work status, n (%)** |  |  |  |  |  |  |
| Employed/self-employed | 40 (70·2) | 26 (74·3) | 14 (63·6) | -10·65 (-35·42, 14·12) | -10·65 (12·64) | 0·39 |
| Employed/self-employed yet not working | 34 (85·0) | 21 (80·8) | 13 (92·9) | 12·09 (-8·20, 32·37) | 12·09 (10·35) | 0·40 |
| Unemployed | 4 (7·0) | 1 (2·9) | 3 (13·6) | 10·78 (-4·59, 26·14) | 10·78 (7·84) | 0·29 |
| Studying | 1 (1·8) | 0 (0·0) | 1 (4·5) | 4·55 (-4·16, 13·25) | 4·55 (4·44) | 0·39 |
| Retired | 7 (12·3) | 6 (17·1) | 1 (4·5) | -12·60 (-27·82, 2·62) | -12·60 (7·77) | 0·23 |
| Other | 5 (8·8) | 2 (5·7) | 3 (13·6) | 7·92 (-8·35, 24·19) | 7·92 (8·30) | 0·36 |
| **Cigarette smoking status, n (%)** |  |  |  |  |  |  |
| Current smoker | 0 (0·0) | 0 (0·0) | 0 (0·0) | - | - | - |
| Previous smoker | 19 (33·3) | 11 (31·4) | 8 (36·4) | 4·94 (-20·37, 30·25) | 4·94 (12·91) | 0·70 |
| Never smoked | 38 (66·7) | 24 (68·6) | 14 (63·6) | -4·94 (-30·25, 20·37) | -4·94 (12·91) | 0·70 |
| **Comorbidities, n (%)** |  |  |  |  |  |  |
| Hypertension | 11 (19·3) | 8 (22·9) | 3 (13·6) | -9·22 (-29·20, 10·76) | -9·22 (10·19) | 0·50 |
| Diabetes | 2 (3·5) | 2 (5·7) | 0 (0·0) | -5·71 (-13·40, 1·98) | -5·71 (3·92) | 0·52 |
| Musculoskeletal disease | 8 (14·0) | 5 (14·3) | 3 (13·6) | -0·65 (-19·09, 17·79) | -0·65 (9·41) | 1·00 |
| Cardiac disease | 4 (7·0) | 0 (0·0) | 4 (18·2) | 18·18 (2·06, 34·30) | 18·18 (8·22) | 0·02* |
| Cancer | 0 (0·0) | 0 (0·0) | 0 (0·0) | - | - | - |
| Respiratory disease (other than COVID) | 10 (17·5) | 5 (14·3) | 5 (22·7) | 8·44 (-12·56, 29·44) | 8·44 (10·72) | 0·41 |
| Number of comorbidities, median (Q1, Q3) | 0·0 (0·0, 1·0) | 0·0 (0·0, 2·0) | 0·0 (0·0, 1·0) | -0·18 (-0·91, 0·54) | -0·18 ± 1·33 | 0·83 |
| Number of comorbidities as category |  |  |  |  |  |  |
| 0 | 31 (54·4) | 19 (54·3) | 12 (54·5) | 0·26 (-26·30, 26·82) | 0·26 (13·55) | 0·98 |
| 1 | 13 (22·8) | 7 (20·0) | 6 (27·3) | 7·27 (-15·57, 30·12) | 7·27 (11·66) | 0·52 |
| 2+ | 13 (22·8) | 9 (25·7) | 4 (18·2) | -7·53 (-29·20, 14·13) | -7·53 (11·05) | 0·75 |
| **Taking prescribed medication, n (%)** | 49 (86·0) | 28 (80·0) | 21 (95·5) | 15·45 (-0·40, 31·31) | 15·45 (8·09) | 0·13 |
| **COVID Frequency, median (Q1, Q3)** | 1·0 (1·0, 2·0) | 1·0 (1·0, 2·0) | 1·0 (1·0, 2·0) | -0·01 (-0·38, 0·36) | -0·01 ± 0·66 | 0·58 |
| **Long covid duration, in months, median (Q1, Q3)** | 14·2 (8·0, 21·4) | 14·8 (6·6, 23·6) | 12·0 (9·1, 21·4) | -0·95 (-6·09, 4·20) | -0·95 ± 9·43 | 0·91 |
| **Number of symptoms, median (Q1, Q3)** | 5·0 (3·0, 7·0) | 4·0 (2·0, 7·0) | 5·0 (4·0, 7·0) | 0·85 (-1·30, 3·00) | 0·85 ± 3·95 | 0·13 |
| **Hospitalization** |  |  |  |  |  |  |
| Hospitalized due to COVID, n (%) |  |  |  |  |  |  |
| 0 | 46 (80·7) | 26 (74·3) | 20 (90·9) | 16·62 (-2·19, 35·44) | 16·62 (9·60) | 0·17 |
| 1 | 8 (14·0) | 7 (20·0) | 1 (4·5) | -15·45 (-31·31, 0·40) | -15·45 (8·09) | 0·13 |
| 2+ | 3 (5·3) | 2 (5·7) | 1 (4·5) | -1·17 (-12·78, 10·45) | -1·17 (5·93) | 1·00 |
| 1st Hospitalization LOS | 3·6 ± 16·5 | 5·8 ± 20·9 | 0·0 ± 0·2 | -5·73 (-11·23, -0·23) | -5·73 ± 16·42 | 0·04* |
| ≥1 ICU admission for COVID, n (%) | 4 (7·0) | 4 (11·4) | 0 (0·0) | -11·43 (-21·97, 0·89) | -11·43 (5·38) | 0·15 |
| 1st ICU LOS | 1·3 ± 6·0 | 2·1 ± 7·5 | 0·0 ± 0·0 | -2·14 (-5·38, 1·09) | -2·14 ± 5·93 | 0·10 |
| **Number of vaccinations, median (Q1, Q3)** | 3·0 (2·0, 4·0) | 4·0 (3·0, 4·0) | 3·0 (2·0, 4·0) | -0·38 (-1·06, 0·29) | -0·38 ± 1·13 | 0·32 |
| **Supplemental oxygen (O<sub>2</sub>), n (%)** | 0 (0·0) | 0 (0·0) | 0 (0·0) | - | - | - |
| **Current level of activity, n (%)** |  |  |  |  |  |  |
| Vigorously active | 0 (0·0) | 0 (0·0) | 0 (0·0) | - | - | - |
| Moderately active | 13 (22·8) | 9 (25·7) | 4 (18·2) | -7·53 (-29·20, 14·13) | -7·53 (11·05) | 0·75 |
| Seldom active | 44 (77·2) | 26 (74·3) | 18 (81·8) | 7·53 (-14·13, 29·20) | 7·53 (11·05) | 0·75 |
| **BDI Total score (0 - 12), median (Q1, Q3)** | 4·0 (2·0, 5·0) | 4·0 (2·0, 6·0) | 3·0 (2·0, 4·0) | -1·15 (-2·42, 0·12) | -1·15 ± 2·33 | 0·16 |
| **DSQ-PEM** |  |  |  |  |  |  |
| PEM (sum of all scores) (0 - 5), median (Q1, Q3) | 4·0 (3·0, 5·0) | 4·0 (2·0, 5·0) | 5·0 (4·0, 5·0) | 0·83 (0·11, 1·55) | 0·83 ± 1·32 | 0·01* |
| PEM score, n (%) |  |  |  |  |  |  |
| 0 | 0 (0·0) | 0 (0·0) | 0 (0·0) | - | - | - |
| 1-2 | 12 (21·1) | 10 (28·6) | 2 (9·1) | -19·48 (-38·67, -0·29) | -19·48 (9·79) | 0·10 |
| 3-5 | 45 (78·9) | 25 (71·4) | 20 (90·9) | 19·48 (0·29, 38·67) | 19·48 (9·79) | 0·10 |
| PEM present with minimal physical efforts, n (%) | 52 (91·2) | 31 (88·6) | 21 (95·5) | 6·88 (-6·79, 20·55) | 6·88 (6·97) | 0·64 |
| PEM present with mental effort, n (%) | 49 (86·0) | 27 (77·1) | 22 (100·0) | 22·86 (8·96, 36·77) | 22·86 (0·00) | 0·02* |

\*p-value < 0·05
Data are presented as mean ± SD unless otherwise specified
AE: Adverse Event; MUHC: McGill University Health Centre; BMI: body mass index; LOS: length of stay; ICU: intensive care unit; BDI: Beck Depression Inventory; DSQ-PEM: DePaul Symptom Questionnaire - post-exertional malaise.

**Table 6 – Supplemental.**
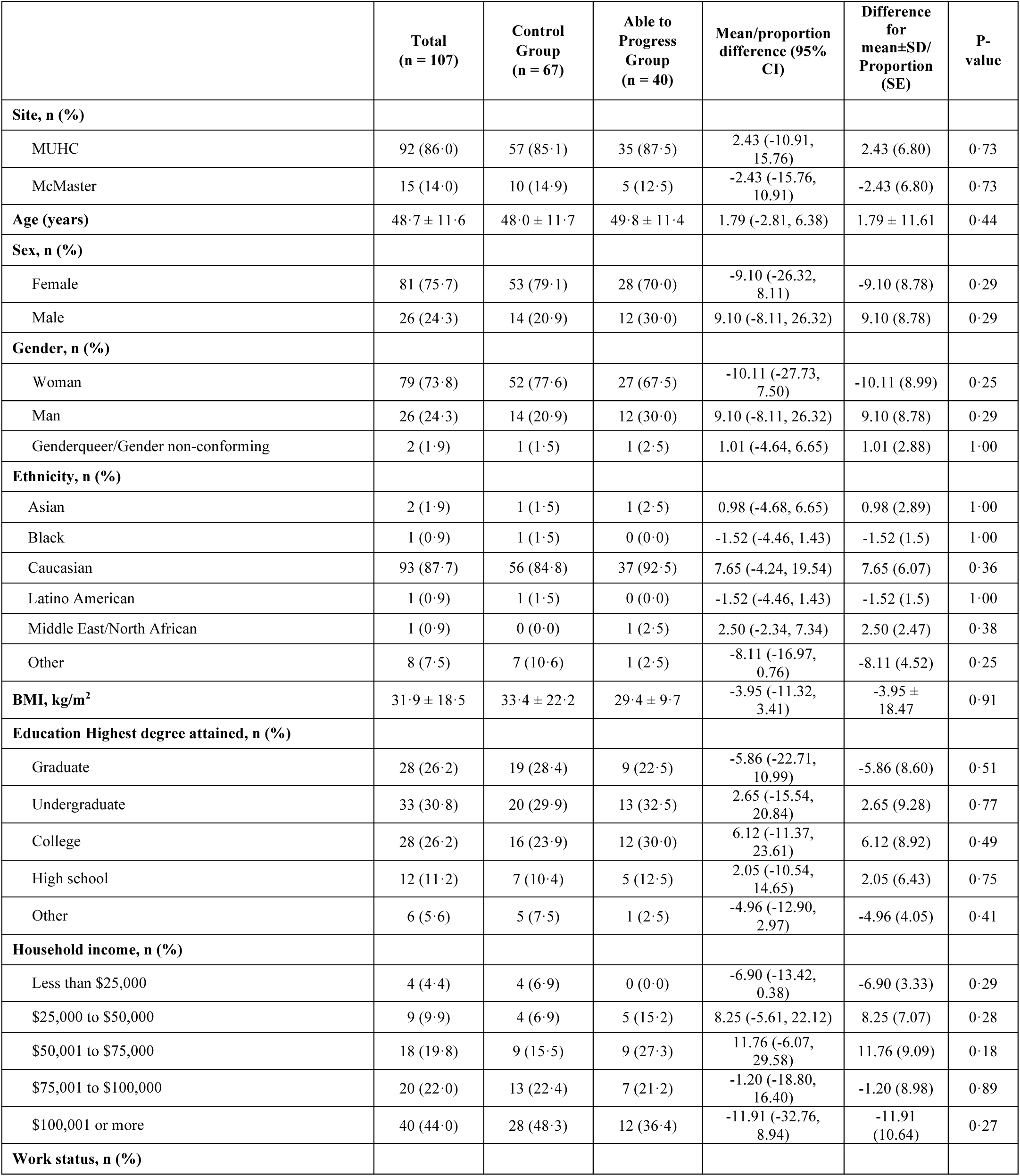
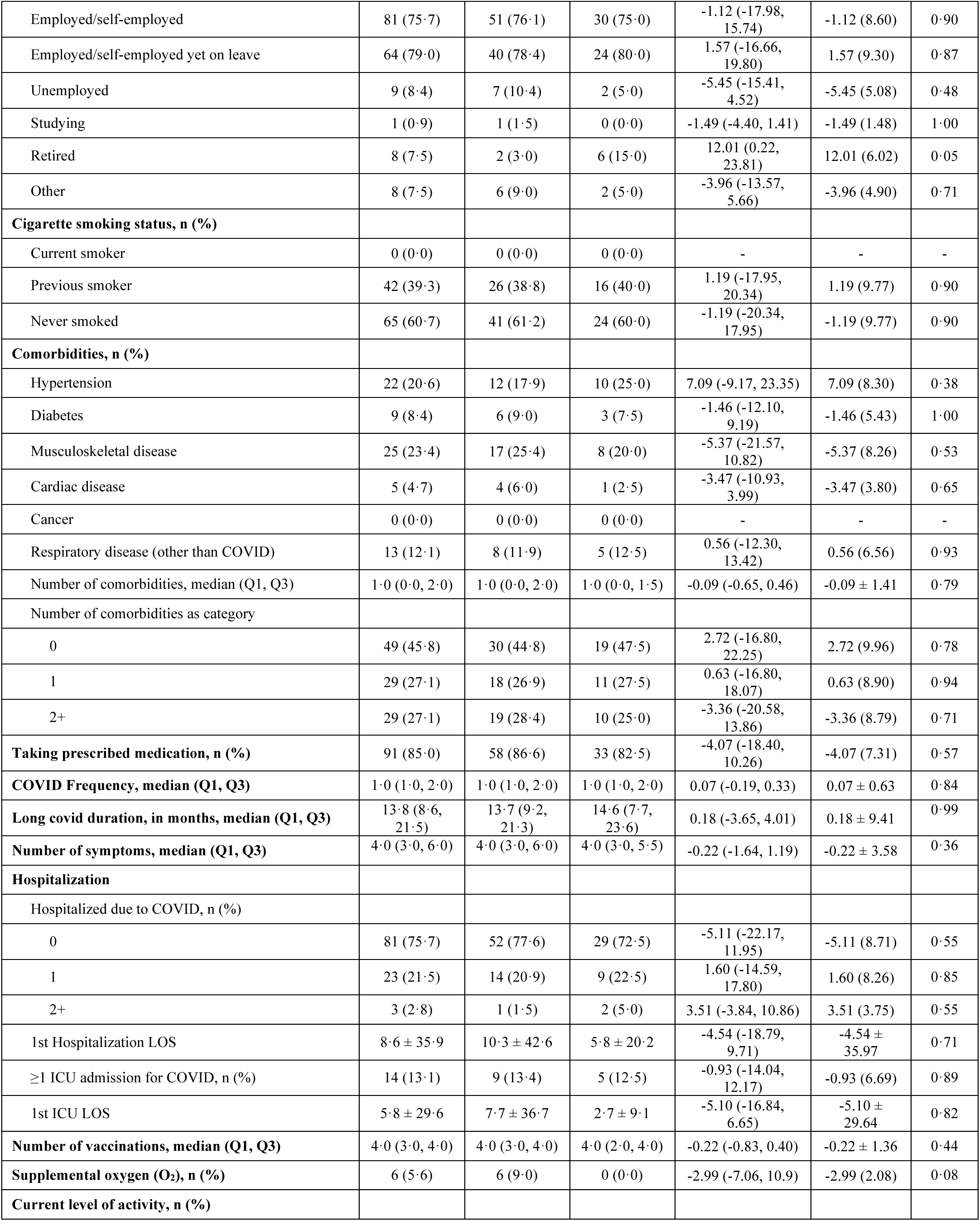
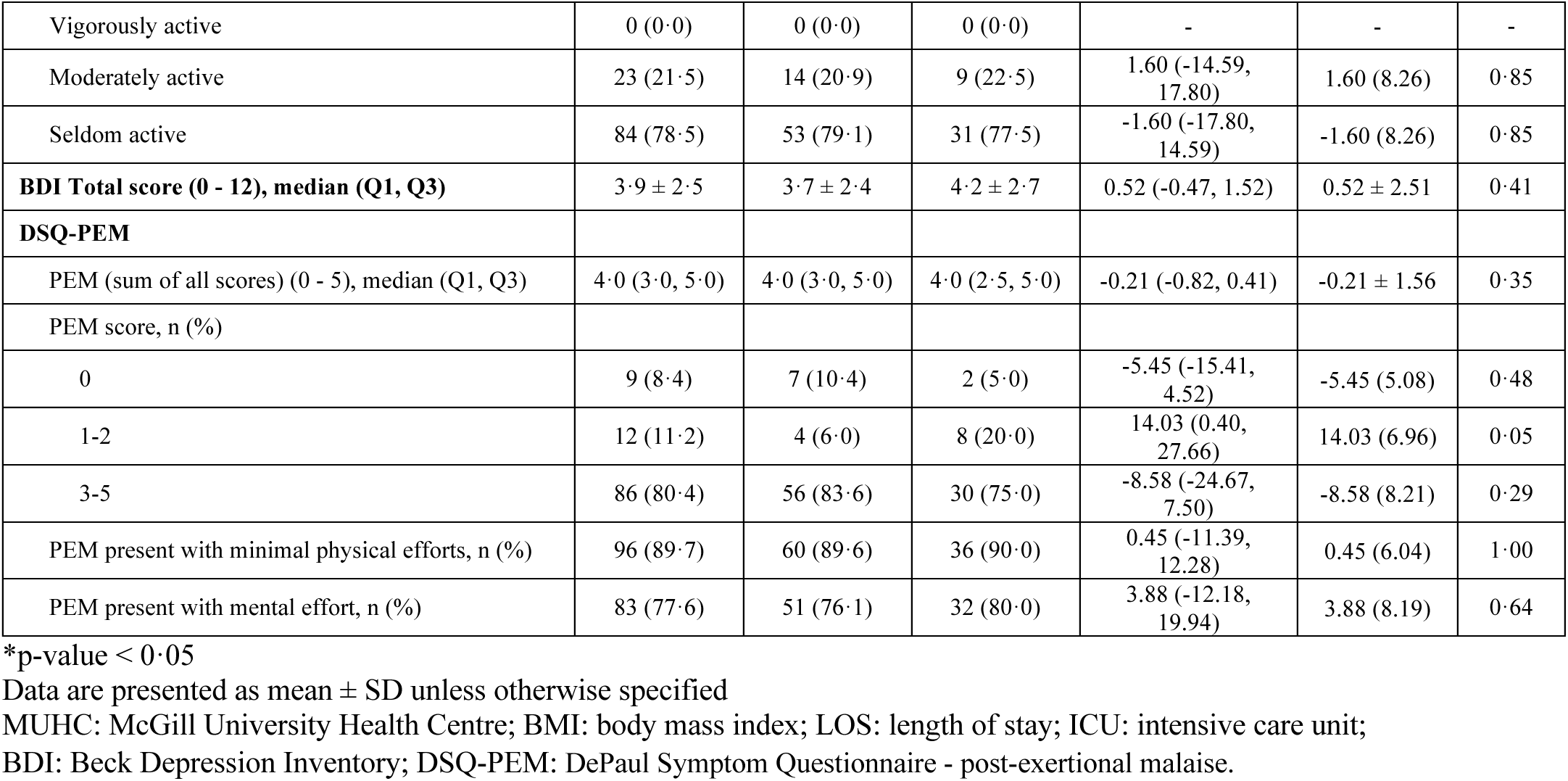
Baseline Characteristics Comparing the Control Group to Participants in the Intervention Group who were Able to Progress Through the Exercise Program.

|  | **Total**<br>**(n = 107)** | **Control**<br>**Group**<br>**(n = 67)** | **Able to**<br>**Progress**<br>**Group**<br>**(n = 40)** | **Mean/proportion**<br>**difference (95%**<br>**CI)** | **Difference**<br>**for**<br>**mean±SD/**<br>**Proportion**<br>**(SE)** | **P-**<br>**value** |
| --- | --- | --- | --- | --- | --- | --- |
| **Site, n (%)** |  |  |  |  |  |  |
| MUHC | 92 (86·0) | 57 (85·1) | 35 (87·5) | 2.43 (-10.91,<br>15.76) | 2.43 (6.80) | 0·73 |
| McMaster | 15 (14·0) | 10 (14·9) | 5 (12·5) | -2.43 (-15.76,<br>10.91) | -2.43 (6.80) | 0·73 |
| **Age (years)** | 48·7 ± 11·6 | 48·0 ± 11·7 | 49·8 ± 11·4 | 1.79 (-2.81, 6.38) | 1.79 ± 11.61 | 0·44 |
| **Sex, n (%)** |  |  |  |  |  |  |
| Female | 81 (75·7) | 53 (79·1) | 28 (70·0) | -9.10 (-26.32,<br>8.11) | -9.10 (8.78) | 0·29 |
| Male | 26 (24·3) | 14 (20·9) | 12 (30·0) | 9.10 (-8.11, 26.32) | 9.10 (8.78) | 0·29 |
| **Gender, n (%)** |  |  |  |  |  |  |
| Woman | 79 (73·8) | 52 (77·6) | 27 (67·5) | -10.11 (-27.73,<br>7.50) | -10.11 (8.99) | 0·25 |
| Man | 26 (24·3) | 14 (20·9) | 12 (30·0) | 9.10 (-8.11, 26.32) | 9.10 (8.78) | 0·29 |
| Genderqueer/Gender non-conforming | 2 (1·9) | 1 (1·5) | 1 (2·5) | 1.01 (-4.64, 6.65) | 1.01 (2.88) | 1·00 |
| **Ethnicity, n (%)** |  |  |  |  |  |  |
| Asian | 2 (1·9) | 1 (1·5) | 1 (2·5) | 0.98 (-4.68, 6.65) | 0.98 (2.89) | 1·00 |
| Black | 1 (0·9) | 1 (1·5) | 0 (0·0) | -1.52 (-4.46, 1.43) | -1.52 (1.5) | 1·00 |
| Caucasian | 93 (87·7) | 56 (84·8) | 37 (92·5) | 7.65 (-4.24, 19.54) | 7.65 (6.07) | 0·36 |
| Latino American | 1 (0·9) | 1 (1·5) | 0 (0·0) | -1.52 (-4.46, 1.43) | -1.52 (1.5) | 1·00 |
| Middle East/North African | 1 (0·9) | 0 (0·0) | 1 (2·5) | 2.50 (-2.34, 7.34) | 2.50 (2.47) | 0·38 |
| Other | 8 (7·5) | 7 (10·6) | 1 (2·5) | -8.11 (-16.97,<br>0.76) | -8.11 (4.52) | 0·25 |
| **BMI, kg/m<sup>2</sup>** | 31·9 ± 18·5 | 33·4 ± 22·2 | 29·4 ± 9·7 | -3.95 (-11.32,<br>3.41) | -3.95 ±<br>18.47 | 0·91 |
| **Education Highest degree attained, n (%)** |  |  |  |  |  |  |
| Graduate | 28 (26·2) | 19 (28·4) | 9 (22·5) | -5.86 (-22.71,<br>10.99) | -5.86 (8.60) | 0·51 |
| Undergraduate | 33 (30·8) | 20 (29·9) | 13 (32·5) | 2.65 (-15.54,<br>20.84) | 2.65 (9.28) | 0·77 |
| College | 28 (26·2) | 16 (23·9) | 12 (30·0) | 6.12 (-11.37,<br>23.61) | 6.12 (8.92) | 0·49 |
| High school | 12 (11·2) | 7 (10·4) | 5 (12·5) | 2.05 (-10.54,<br>14.65) | 2.05 (6.43) | 0·75 |
| Other | 6 (5·6) | 5 (7·5) | 1 (2·5) | -4.96 (-12.90,<br>2.97) | -4.96 (4.05) | 0·41 |
| **Household income, n (%)** |  |  |  |  |  |  |
| Less than \$25,000 | 4 (4·4) | 4 (6·9) | 0 (0·0) | -6.90 (-13.42,<br>0.38) | -6.90 (3.33) | 0·29 |
| \$25,000 to \$50,000 | 9 (9·9) | 4 (6·9) | 5 (15·2) | 8.25 (-5.61, 22.12) | 8.25 (7.07) | 0·28 |
| \$50,001 to \$75,000 | 18 (19·8) | 9 (15·5) | 9 (27·3) | 11.76 (-6.07,<br>29.58) | 11.76 (9.09) | 0·18 |
| \$75,001 to \$100,000 | 20 (22·0) | 13 (22·4) | 7 (21·2) | -1.20 (-18.80,<br>16.40) | -1.20 (8.98) | 0·89 |
| \$100,001 or more | 40 (44·0) | 28 (48·3) | 12 (36·4) | -11.91 (-32.76,<br>8.94) | -11.91<br>(10.64) | 0·27 |
| **Work status, n (%)** |  |  |  |  |  |  |
| Employed/self-employed | 81 (75·7) | 51 (76·1) | 30 (75·0) | -1.12 (-17.98, 15.74) | -1.12 (8.60) | 0·90 |
| Employed/self-employed yet on leave | 64 (79·0) | 40 (78·4) | 24 (80·0) | 1.57 (-16.66, 19.80) | 1.57 (9.30) | 0·87 |
| Unemployed | 9 (8·4) | 7 (10·4) | 2 (5·0) | -5.45 (-15.41, 4.52) | -5.45 (5.08) | 0·48 |
| Studying | 1 (0·9) | 1 (1·5) | 0 (0·0) | -1.49 (-4.40, 1.41) | -1.49 (1.48) | 1·00 |
| Retired | 8 (7·5) | 2 (3·0) | 6 (15·0) | 12.01 (0.22, 23.81) | 12.01 (6.02) | 0·05 |
| Other | 8 (7·5) | 6 (9·0) | 2 (5·0) | -3.96 (-13.57, 5.66) | -3.96 (4.90) | 0·71 |
| **Cigarette smoking status, n (%)** |  |  |  |  |  |  |
| Current smoker | 0 (0·0) | 0 (0·0) | 0 (0·0) | - | - | - |
| Previous smoker | 42 (39·3) | 26 (38·8) | 16 (40·0) | 1.19 (-17.95, 20.34) | 1.19 (9.77) | 0·90 |
| Never smoked | 65 (60·7) | 41 (61·2) | 24 (60·0) | -1.19 (-20.34, 17.95) | -1.19 (9.77) | 0·90 |
| **Comorbidities, n (%)** |  |  |  |  |  |  |
| Hypertension | 22 (20·6) | 12 (17·9) | 10 (25·0) | 7.09 (-9.17, 23.35) | 7.09 (8.30) | 0·38 |
| Diabetes | 9 (8·4) | 6 (9·0) | 3 (7·5) | -1.46 (-12.10, 9.19) | -1.46 (5.43) | 1·00 |
| Musculoskeletal disease | 25 (23·4) | 17 (25·4) | 8 (20·0) | -5.37 (-21.57, 10.82) | -5.37 (8.26) | 0·53 |
| Cardiac disease | 5 (4·7) | 4 (6·0) | 1 (2·5) | -3.47 (-10.93, 3.99) | -3.47 (3.80) | 0·65 |
| Cancer | 0 (0·0) | 0 (0·0) | 0 (0·0) | - | - | - |
| Respiratory disease (other than COVID) | 13 (12·1) | 8 (11·9) | 5 (12·5) | 0.56 (-12.30, 13.42) | 0.56 (6.56) | 0·93 |
| Number of comorbidities, median (Q1, Q3) | 1·0 (0·0, 2·0) | 1·0 (0·0, 2·0) | 1·0 (0·0, 1·5) | -0.09 (-0.65, 0.46) | -0.09 ± 1.41 | 0·79 |
| Number of comorbidities as category |  |  |  |  |  |  |
| 0 | 49 (45·8) | 30 (44·8) | 19 (47·5) | 2.72 (-16.80, 22.25) | 2.72 (9.96) | 0·78 |
| 1 | 29 (27·1) | 18 (26·9) | 11 (27·5) | 0.63 (-16.80, 18.07) | 0.63 (8.90) | 0·94 |
| 2+ | 29 (27·1) | 19 (28·4) | 10 (25·0) | -3.36 (-20.58, 13.86) | -3.36 (8.79) | 0·71 |
| **Taking prescribed medication, n (%)** | 91 (85·0) | 58 (86·6) | 33 (82·5) | -4.07 (-18.40, 10.26) | -4.07 (7.31) | 0·57 |
| **COVID Frequency, median (Q1, Q3)** | 1·0 (1·0, 2·0) | 1·0 (1·0, 2·0) | 1·0 (1·0, 2·0) | 0.07 (-0.19, 0.33) | 0.07 ± 0.63 | 0·84 |
| **Long covid duration, in months, median (Q1, Q3)** | 13·8 (8·6, 21·5) | 13·7 (9·2, 21·3) | 14·6 (7·7, 23·6) | 0.18 (-3.65, 4.01) | 0.18 ± 9.41 | 0·99 |
| **Number of symptoms, median (Q1, Q3)** | 4·0 (3·0, 6·0) | 4·0 (3·0, 6·0) | 4·0 (3·0, 5·5) | -0.22 (-1.64, 1.19) | -0.22 ± 3.58 | 0·36 |
| **Hospitalization** |  |  |  |  |  |  |
| Hospitalized due to COVID, n (%) |  |  |  |  |  |  |
| 0 | 81 (75·7) | 52 (77·6) | 29 (72·5) | -5.11 (-22.17, 11.95) | -5.11 (8.71) | 0·55 |
| 1 | 23 (21·5) | 14 (20·9) | 9 (22·5) | 1.60 (-14.59, 17.80) | 1.60 (8.26) | 0·85 |
| 2+ | 3 (2·8) | 1 (1·5) | 2 (5·0) | 3.51 (-3.84, 10.86) | 3.51 (3.75) | 0·55 |
| 1st Hospitalization LOS | 8·6 ± 35·9 | 10·3 ± 42·6 | 5·8 ± 20·2 | -4.54 (-18.79, 9.71) | -4.54 ± 35.97 | 0·71 |
| ≥1 ICU admission for COVID, n (%) | 14 (13·1) | 9 (13·4) | 5 (12·5) | -0.93 (-14.04, 12.17) | -0.93 (6.69) | 0·89 |
| 1st ICU LOS | 5·8 ± 29·6 | 7·7 ± 36·7 | 2·7 ± 9·1 | -5.10 (-16.84, 6.65) | -5.10 ± 29.64 | 0·82 |
| **Number of vaccinations, median (Q1, Q3)** | 4·0 (3·0, 4·0) | 4·0 (3·0, 4·0) | 4·0 (2·0, 4·0) | -0.22 (-0.83, 0.40) | -0.22 ± 1.36 | 0·44 |
| **Supplemental oxygen (O<sub>2</sub>), n (%)** | 6 (5·6) | 6 (9·0) | 0 (0·0) | -2.99 (-7.06, 10.9) | -2.99 (2.08) | 0·08 |
| **Current level of activity, n (%)** |  |  |  |  |  |  |
| Vigorously active | 0 (0·0) | 0 (0·0) | 0 (0·0) | - | - | - |
| Moderately active | 23 (21·5) | 14 (20·9) | 9 (22·5) | 1.60 (-14.59, 17.80) | 1.60 (8.26) | 0·85 |
| Seldom active | 84 (78·5) | 53 (79·1) | 31 (77·5) | -1.60 (-17.80, 14.59) | -1.60 (8.26) | 0·85 |
| **BDI Total score (0 - 12), median (Q1, Q3)** | 3·9 ± 2·5 | 3·7 ± 2·4 | 4·2 ± 2·7 | 0.52 (-0.47, 1.52) | 0.52 ± 2.51 | 0·41 |
| **DSQ-PEM** |  |  |  |  |  |  |
| PEM (sum of all scores) (0 - 5), median (Q1, Q3) | 4·0 (3·0, 5·0) | 4·0 (3·0, 5·0) | 4·0 (2·5, 5·0) | -0.21 (-0.82, 0.41) | -0.21 ± 1.56 | 0·35 |
| PEM score, n (%) |  |  |  |  |  |  |
| 0 | 9 (8·4) | 7 (10·4) | 2 (5·0) | -5.45 (-15.41, 4.52) | -5.45 (5.08) | 0·48 |
| 1-2 | 12 (11·2) | 4 (6·0) | 8 (20·0) | 14.03 (0.40, 27.66) | 14.03 (6.96) | 0·05 |
| 3-5 | 86 (80·4) | 56 (83·6) | 30 (75·0) | -8.58 (-24.67, 7.50) | -8.58 (8.21) | 0·29 |
| PEM present with minimal physical efforts, n (%) | 96 (89·7) | 60 (89·6) | 36 (90·0) | 0.45 (-11.39, 12.28) | 0.45 (6.04) | 1·00 |
| PEM present with mental effort, n (%) | 83 (77·6) | 51 (76·1) | 32 (80·0) | 3.88 (-12.18, 19.94) | 3.88 (8.19) | 0·64 |

\*p-value < 0·05
Data are presented as mean ± SD unless otherwise specified
MUHC: McGill University Health Centre; BMI: body mass index; LOS: length of stay; ICU: intensive care unit;
BDI: Beck Depression Inventory; DSQ-PEM: DePaul Symptom Questionnaire - post-exertional malaise.

**Table 7 – Supplemental.** Per-Protocol Analysis Comparing the Control Group to Participants in the Intervention Group who were Able to Progress Through the Exercise Program.

|  | Control Group<br>(n = 66) |  | Able to Progress<br>Group<br>(n = 36) |  | Univariate Analysis |  |
| --- | --- | --- | --- | --- | --- | --- |
|  | Change post-intervention<br>(Post-Pre) |  | Change post-intervention<br>(Post-Pre) |  | Difference in change post-intervention (Post-Pre) | Comparing<br>the change in<br>the two arms |
|  | mean/proportion (95% CI) | P-value | mean/proportion (95% CI) | P-value | mean/proportion (95% CI) | P-value |
| **AM-PAC** |  |  |  |  |  |  |
| Mobility T-Score | 1.0 (0.0, 1.9) | 0.04* | 2.4 (0.9, 3.9) | 0.00* | 1.44 (-0.37, 3.25) | 0.11 |
| Cognitive T-Score | 1.6 (0.1, 3.1) | 0.04* | 3.3 (1.2, 5.4) | 0.00* | 1.71 (-0.83, 4.25) | 0.18 |
| **1-min STS** |  |  |  |  |  |  |
| Number of repetitions in 1 minute | 3.2 (2.0, 4.3) | <0.001* | 4.8 (3.4, 6.2) | <0.001* | 1.60 (-0.18, 3.39) | 0.06 |
| **TUG** |  |  |  |  |  |  |
| Time to complete TUG (seconds) | -0.8 (-1.5, -0.1) | 0.03* | -0.8 (-1.5, -0.1) | 0.02* | -0.08 (-1.06, 0.90) | 0.88 |
| **SF-12** |  |  |  |  |  |  |
| Mental Score (0 - 100) | 3.6 (0.9, 6.3) | 0.01* | 7.4 (3.9, 10.9) | <0.001* | 3.78 (-0.69, 8.25) | 0.10 |
| Physical Score (0 - 100) | 0.2 (-1.9, 2.3) | 0.86 | 4.2 (1.5, 7.0) | 0.00* | 4.04 (0.61, 7.46) | 0.02* |
| **EQ-5D-5L** |  |  |  |  |  |  |
| Mobility (1 - 5) | -0.0 (-0.3, 0.2) | 0.70 | -0.3 (-0.7, -0.0) | 0.04* | -0.29 (-0.69, 0.11) | 0.15 |
| Self-care (1 - 5) | 0.2 (-0.0, 0.3) | 0.09 | -0.1 (-0.3, 0.2) | 0.52 | -0.23 (-0.52, 0.06) | 0.12 |
| Usual Activities (1 - 5) | -0.2 (-0.5, 0.0) | 0.06 | -0.4 (-0.7, 0.0) | 0.06 | -0.13 (-0.57, 0.31) | 0.56 |
| Pain / Discomfort (1 - 5) | -0.0 (-0.2, 0.2) | 0.88 | -0.5 (-0.8, -0.1) | 0.00* | -0.45 (-0.84, -0.06) | 0.03* |
| Anxiety / Depression (1 - 5) | -0.1 (-0.3, 0.1) | 0.50 | -0.3 (-0.6, 0.0) | 0.09 | -0.23 (-0.64, 0.19) | 0.28 |
| VAS (0 - 100) | 2.5 (-1.9, 7.0) | 0.27 | 14.0 (7.1, 20.9) | <0.001* | 11.48 (3.24, 19.73) | 0.01* |
| **VAS fatigue (0 - 10)** | -0.8 (-1.4, -0.3) | 0.00* | -2.2 (-3.1, -1.4) | <0.001* | -1.40 (-2.42, -0.39) | 0.01* |
| **HADS - anxiety score (0 - 21)** | -0.2 (-1.0, 0.6) | 0.69 | -1.5 (-2.5, -0.5) | 0.00* | -1.38 (-2.68, -0.08) | 0.04* |
| Borderline (8 - 10), proportion % (SE) | 1.2 (-11.6, 14.0) | 0.86 | -1.2 (-13.0, 10.5) | 0.84 | -2.44 (-19.97, 15.09) | 0.79 |
| Abnormal (11 - 21), proportion % (SE) | 1.3 (-9.3, 12.0) | 0.81 | -10.5 (-22.9, 1.9) | 0.10 | -11.84 (-28.24, 4.55) | 0.16 |
| **HADS - depression score (0 - 21)** | -0.8 (-1.8, 0.1) | 0.07 | -2.1 (-3.3, -0.9) | <0.001* | -1.22 (-2.74, 0.29) | 0.11 |
| Borderline (8 - 10), proportion % (SE) | -2.4 (-15.7, 10.9) | 0.73 | -16.2 (-32.5, -0.0) | 0.05* | -13.86 (-34.85, 7.13) | 0.20 |
| Abnormal (11 - 21), proportion % (SE) | -3.9 (-14.7, 6.9) | 0.48 | -9.5 (-23.8, 4.8) | 0.19 | -5.62 (-23.65, 12.41) | 0.54 |
| **IES-R score** | -4.9 (-8.8, -1.0) | 0.02* | -2.3 (-7.0, 2.5) | 0.35 | 2.61 (-3.43, 8.65) | 0.40 |
| ≥33, proportion % (SE) | -2.2 (-13.6, 9.1) | 0.70 | 1.0 (-14.0, 16.0) | 0.90 | 3.24 (-15.34, 21.82) | 0.73 |
| ≥37, proportion % (SE) | -5.4 (-16.6, 5.9) | 0.35 | 3.0 (-9.2, 15.2) | 0.63 | 8.37 (-8.19, 24.94) | 0.32 |
| **DSQ-PEM** | **n = 34** |  | **n = 23** |  |  |  |
| PEM (sum of all scores) (0 - 5) | -1.5 (-2.2, -0.8) | <0.001* | -2.5 (-3.3, -1.7) | <0.001* | -0.98 (-2.05, 0.10) | 0.08 |
| PEM present with minimal physical effort, proportion % (SE) | -25.1 (-43.6, -6.6) | 0.01* | -49.6 (-69.5, -29.7) | <0.001* | -24.51 (-51.39, 2.37) | 0.07 |
| PEM present with mental effort, proportion % (SE) | -8.8 (-28.4, 10.8) | 0.38 | -8.9 (-26.3, 8.5) | 0.32 | -0.11 (-26.87, 26.65) | 0.99 |
|  | **Control Group**<br>**(n = 66)** |  | **Intervention Group**<br>**(n = 36)** |  | **Difference between the two arms** | **P-value** |
|  | **Post-intervention** |  | **Post-intervention** |  | **mean/proportion (95% CI)** | **Comparing the two arms** |
| **TDI** |  |  |  |  |  |  |
| Functional score, mean ± SD | 0.4 ± 0.8 |  | 1.0 ± 0.9 |  | 0.67 (0.32, 1.02) | <0.001* |
| Magnitude score, mean ± SD | 0.6 ± 1.3 |  | 1.0 ± 1.4 |  | 0.40 (-0.14, 0.95) | 0.06 |
| Effort score, mean ± SD | 0.5 ± 1.4 |  | 1.6 ± 1.3 |  | 1.05 (0.51, 1.59) | <0.001* |
| Total score, mean ± SD | 1.5 ± 3.0 |  | 3.6 ± 3.0 |  | 2.11 (0.92, 3.31) | <0.001* |
| **HCU** |  |  |  |  |  |  |
| Doctor visit, n (%) | 30 (46.2) |  | 16 (44.4) |  | -1.71 (-21.97, 18.55) | 1.00 |
| ER visit, n (%) | 3 (4.6) |  | 0 (0.0) |  | -4.62 (-9.72, 0.49) | 0.55 |
| Hospitalization, n (%) | 1 (1.5) |  | 0 (0.0) |  | -1.54 (-4.53, 1.45) | 1.00 |

\*p-value < 0.05
1-min STS: 1-minute sit-to-stand test; SE: standard error; SpO<sub>2</sub>: saturation of peripheral oxygen; HR: heart rate; TUG: Timed-Up-and-Go test; AM-PAC: Activity Measure for Post-Acute Care; SF-12: health-related quality of life questionnaire; 5Q-5D-5L: EuroQol 5 Dimension 5 Level; VAS: visual analogue scale; HADS: Hospital Anxiety and Depression scale; IES-R: Impact of Event Scale – Revised; DSQ-PEM: DePaul Symptom Questionnaire - post-exertional malaise; TDI: Transition Dyspnea Index; HCU: Health Care Utilization.
IES-R score: ≥ 33 represents the best cutoff for a probable diagnosis of PTSD, ≥ 37 high enough to suppress immune system functioning.

**Table 8 – Supplemental.** Between-Group Analyses Stratified by Sex.

|  | Female (n = 99) |  | Men (n = 33) |  |
| --- | --- | --- | --- | --- |
|  | Difference in change post-intervention (Post-Pre) | Comparing the change in the two arms | Difference in change post-intervention (Post-Pre) | Comparing the change in the two arms |
|  | mean/proportion (95% CI) | P-value | mean/proportion (95% CI) | P-value |
| **AM-PAC** |  |  |  |  |
| Mobility T-Score | 1.18 (-0.45, 2.81) | 0.15 | -1.45 (-4.93, 2.04) | 0.41 |
| Cognitive T-Score | 1.00 (-1.35, 3.36) | 0.40 | 2.06 (-2.22, 6.35) | 0.34 |
| **1-min STS** |  |  |  |  |
| Number of repetitions in 1 minute | 0.44 (-1.54, 2.42) | 0.66 | -0.97 (-4.31, 2.38) | 0.57 |
| **TUG** |  |  |  |  |
| Time to complete TUG (seconds) | 0.43 (-0.54, 1.41) | 0.38 | 1.22 (-1.39, 3.83) | 0.36 |
| **SF-12** |  |  |  |  |
| Mental Score (0 - 100) | 3.12 (-1.08, 7.31) | 0.15 | -1.16 (-9.08, 6.77) | 0.78 |
| Physical Score (0 - 100) | 2.77 (-0.58, 6.13) | 0.11 | 0.88 (-5.07, 6.83) | 0.77 |
| **EQ-5D-5L** |  |  |  |  |
| Mobility (1 - 5) | -0.29 (-0.67, 0.08) | 0.13 | -0.15 (-0.81, 0.50) | 0.65 |
| Self-care (1 - 5) | -0.11 (-0.42, 0.20) | 0.50 | -0.20 (-0.71, 0.32) | 0.46 |
| Usual Activities (1 - 5) | 0.09 (-0.32, 0.50) | 0.68 | -0.23 (-0.97, 0.51) | 0.54 |
| Pain / Discomfort (1 - 5) | -0.36 (-0.73, 0.01) | 0.06 | -0.30 (-1.06, 0.46) | 0.44 |
| Anxiety / Depression (1 - 5) | -0.17 (-0.54, 0.19) | 0.35 | -0.04 (-0.83, 0.74) | 0.91 |
| VAS (0 - 100) | 8.15 (0.50, 15.79) | 0.04* | 5.88 (-8.48, 20.25) | 0.42 |
| **VAS fatigue (0 - 10)** | -0.61 (-1.57, 0.36) | 0.22 | -1.35 (-3.05, 0.34) | 0.12 |
| **HADS - anxiety score (0 - 21)** | -0.88 (-2.10, 0.33) | 0.15 | -0.76 (-3.35, 1.84) | 0.57 |
| Borderline (8 - 10), proportion % (SE) | -0.77 (-27.83, 26.30) | 0.96 | -7.11 (-57.83, 43.61) | 0.78 |
| Abnormal (11 - 21), proportion % (SE) | -3.34 (-28.64, 21.96) | 0.80 | -6.54 (-29.51, 16.43) | 0.58 |
| **HADS - depression score (0 - 21)** | -0.49 (-2.09, 1.11) | 0.55 | -1.07 (-3.68, 1.54) | 0.42 |
| Borderline (8 - 10), proportion % (SE) | -5.11 (-30.22, 20.00) | 0.69 | -11.29 (-61.26, 38.68) | 0.66 |
| Abnormal (11 - 21), proportion % (SE) | 0·18 (-26·24, 26·60) | 0·99 | -10·12 (-58·17, 37·92) | 0·68 |
| **IES-R score** | 6·46 (-0·13, 13·06) | 0·06 | -2·03 (-10·72, 6·66) | 0·65 |
| ≥33, proportion % (SE) | -0·67 (-21·85, 20·50) | 0·95 | 7·11 (-37·57, 51·79) | 0·76 |
| ≥37, proportion % (SE) | 6·32 (-13·41, 26·06) | 0·53 | 6·40 (-38·74, 51·55) | 0·78 |
| **DSQ-PEM** | **n = 53** |  | **n = 17** |  |
| PEM (sum of all scores) (0 - 5) | -0·44 (-1·60, 0·72) | 0·45 | -0·08 (-1·80, 1·63) | 0·92 |
| PEM present with minimal physical effort, proportion % (SE) | -2·91 (-31·38, 25·55) | 0·84 | -16·35 (-50·22, 17·53) | 0·34 |
| PEM present with mental effort , proportion % (SE) | 1·68 (-34·85, 38·21) | 0·93 | -22·23 (-69·63, 25·16) | 0·36 |
|  | **Female**<br>**(n = 99)** | **Male**<br>**(n = 33)** | **Difference between**<br>**male and female** | **P-value** |
|  | **Post-intervention** | **Post-intervention** | **mean/proportion**<br>**(95% CI)** | **Comparing**<br>**male and**<br>**female** |
| **TDI** |  |  |  |  |
| Functional score, mean ± SD | 0·5 ± 1·0 | 0·6 ± 0·8 | 0·08 (-0·30, 0·46) | 0·67 |
| Magnitude score, mean ± SD | 0·7 ± 1·3 | 0·9 ± 1·6 | 0·23 (-0·33, 0·80) | 0·31 |
| Effort score, mean ± SD | 0·8 ± 1·5 | 0·8 ± 1·5 | -0·06 (-0·66, 0·54) | 0·93 |
| Total score, mean ± SD | 2·0 ± 3·3 | 2·3 ± 3·3 | 0·29 (-1·05, 1·63) | 0·52 |
| **HCU** |  |  |  |  |
| Doctor visit, n (%) | 52 (55·3) | 12 (37·5) | -17·82 (-37·37, 1·74) | 0·10 |
| ER visit, n (%) | 3 (3·2) | 1 (3·1) | -0·07 (-7·06, 6·93) | 1·00 |
| Hospitalization, n (%) | 2 (2·1) | 0 (0·0) | -2·13 (-0·79, 5·04) | 1·00 |

\*p-value < 0·05
1-min STS: 1-minute sit-to-stand test; SE: standard error; SpO<sub>2</sub>: saturation of peripheral oxygen; HR: heart rate; TUG: Timed-Up-and-Go test; AM-PAC: Activity Measure for Post-Acute Care; SF-12: health-related quality of life questionnaire; 5Q-5D-5L: EuroQol 5 Dimension 5 Level; VAS: visual analogue scale; HADS: Hospital Anxiety and Depression scale; IES-R: Impact of Event Scale – Revised; DSQ-PEM: DePaul Symptom Questionnaire - post-exertional malaise; TDI: Transition Dyspnea Index; HCU: Health Care Utilization.
IES-R score: ≥ 33 represents the best cutoff for a probable diagnosis of PTSD, ≥ 37 high enough to suppress immune system functioning.

